# Vasopressin and angiotensin II differentially modulate human fear response dynamics to looming threats

**DOI:** 10.1101/2025.05.18.25327885

**Authors:** Mengfan Han, Wenyi Dong, Kun Fu, Junjie Wang, Yuanhang Xu, Yueyuan Zheng, Keith M Kendrick, Ferraro Stefania, Ting Xu, Dezhong Yao, Benjamin Becker

## Abstract

While basal threat processing dynamics (e.g., visual looming) are well characterized in animals, the underlying mechanisms and their modulation by neuropeptide systems with different modulatory roles in threat processing (vasopressin, angiotensin II) remain poorly understood in humans. In a randomized, placebo-controlled eye-tracking study (*N*=111), we administered vasopressin (AVP) or an angiotensin II receptor blocker (via losartan, LT) during a time-to-collision threat paradigm. Behaviorally, AVP induced a systematic time overestimation while LT induced temporal compression and reduced state anxiety. Pupillometry revealed distinguishable profiles: AVP induced sustained constriction during stimulus approach followed by post-stimulus threat-specific dilation, LT maintained sustained pupillary constriction throughout both approach and occlusion phases yet preserving threat-specificity, while placebo (PLC) showed no threat-specific modulation. A computational framework (combining functional principal component analysis [FPCA], clustering, and hidden Markov model [HMM]) underscored the distinct modulations: AVP stabilized a high-arousal state characterized by the co-activation of vigilance, threat-proactive preparation and a shift from perception to internal simulation. LT suppressed transitions to high-arousal states and exhibited maximal sequence entropy, reflecting flexible response patterns—contrasting with placebo’s lowest entropy dynamics. These results demonstrate that AVP and LT differentially regulate basal threat processing via separable neuropeptide pathways: AVP sustains hypervigilance while LT promotes anxiolysis and adaptive flexibility. Our findings suggest neuropeptide pathway-specific targets maladaptive threat processing in trauma- or anxiety-related disorders.

## Introduction

The subjective experience of emotions varies across individuals and requires the coordinated involvement of distributed cortical and subcortical systems [1–3]. Some basic forms of ‘primitive’ affective mechanisms such as defensive reactions to rapidly approaching objects (‘looming’) appear to be innate, highly preserved across phylogenetic and ontogenetic evolution and tightly linked to specific (subcortical) circuits [4,5]. The cognitive and neural bases of the corresponding affective processes have been extensively studied within broader models of emotion. Foundational work has highlighted the central role of subcortical structures, including the amygdala, in the rapid and automatic detection of motivationally salient stimuli, particularly threats [6,7]. This rapid threat detection system interacts with cortical networks to generate conscious emotional experiences and coordinate adaptive behavioral responses [8].

In interaction with these foundational mechanisms, the visual system can rapidly detect impending – and potentially threatening - collisions via estimating the accelerating expansion of objects on the retina (looming). Looming presents a fundamental, evolutionary and highly conserved threat detection mechanisms, which can be observed from fruit flies to humans [9,10]. This innate perceptual sensitivity to approaching stimuli that specifies the time-to-collision independent of size or distance enables avoiding collisions and rapid defensive responses [11–13].

Although looming perception has been traditionally viewed as a basic visual computation, recent studies have demonstrated that looming is modulated by the semantic and affective content of the stimuli, such that threatening stimuli are typically underestimated in the time to collision by 20-30%, a time compression effect that may critically facilitate faster defensive reactions [13–15]. However, in natural environments, threatening objects such as predators often temporarily disappear from view, raising critical questions about how threat processing mechanisms operate without continuous visual input and how imagination maintains threat assessment during such periods. Understanding these mechanisms is crucial for developing ecologically valid models of threat processing and may help to determine how these processes can be targeted in mental disorders characterized by excessive threat processing.

The neuropeptide systems arginine vasopressin (AVP) and renin-angiotensin (RAS) have been associated with threat-related processing across species. AVP, synthesized in the hypothalamus, enhances threat processing by increasing the extended amygdala sensitivity to threat information and modulating prefrontal-amygdala circuits [16,17], with pronounced effects in males [18]. The RAS, in particular blockade of the RAS II AT1 receptors, has been associated with decreased fear by reducing amygdala reactivity to threat and enhancing prefrontal regulatory control [5,19–21]. While both systems influence threat processing through partially overlapping neural circuits and show complex synergistic effects in regulating physiological indices of threat responsivity including blood pressure and stress responses [22–24] their respective roles in looming threat processing remain to be explored.

Pupillary responses offer unique advantages as precise and non-invasive indicators of threat processing dynamics [25–27]. Through tracking the activity of the locus coeruleus-norepinephrine (LC-NE) system pupil dilation provides high temporal resolution information about arousal related to cognitive and affective engagement [28,29], As such, pupil responses can reveal the continuous temporal dynamics of threat processing.

The LC-NE arousal system has shown a high sensitivity to both AVP [30] and RAS [31] modulation which render pupillary responses an ideal indicator for investigating the neurobiological basis of their modulatory role on threat processing.

Against this background we aimed at determining the role of two neuropeptide systems (AVP, RAS) in basal threat-related mechanisms, in particular to determine their effects on threat processing during looming via determining the effects of single dosages of both, AVP and a selective and competitive angiotensin receptor II antagonists (Losartan, LT) on behavioral and pupillary responses during the looming fear paradigm in combination with computational analytic approaches. This novel combination allows us to dissect the distinct contributions of these systems to threat processing and explore potential therapeutic implications. The hypothesized effects were tested in a pre-registered randomized placebo-controlled double-blind psychopharmacological eye tracking design during which participants were administered oral AVP, LT or placebo (PLC).

To provide an accurate test of the complex hypotheses we established a novel expansion of the threat looming paradigm and established a comprehensive analytic framework. This expansion integrates both visual and imagination components, comprising two phases: a one-second ’visual stimulus amplification’ phase providing initial approach velocity information, followed by an ’imagined stimulus approach’ phase where the visual threat stimulus disappears after the approach information has been acquired. This design builds on substantial evidence that threat-related neural systems remain active even without direct visual input [8,32], and amygdala-centered defensive circuits continue processing threats when visual stimuli are subliminal or masked [33,34], indicating robust threat maintenance mechanisms during temporary visual occlusion.

To characterize the complex dynamics of threat processing, we developed an integrated analytical framework combining pupillary features, behavioral responses, and temporal measurements. This multivariate approach identifies distinct threat response patterns and their pharmacological modulation through dimensional reduction and clustering techniques. By simultaneously analyzing multiple pupillary characteristics and behavioral measures, we reveal how different pharmacological treatments shape overall threat processing strategies. This systematic approach advances beyond traditional univariate analyses to capture the temporal evolution and multidimensional nature of threat responses under different pharmacological conditions.

Our specific aims in this study were to: (1) characterize threat processing dynamics during looming through pupillary responses, (2) examine the maintenance of threat perception during visual occlusion, (3) investigate pharmacological modulation of the identified threat processes, and (4) identify distinct response patterns through Functional Principal Component Analysis–clustering–Hidden Markov Model (FPCA–clustering–HMM) analysis. The findings may reveal innovative targets for testing novel interventions for disorders characterized by elevated threat reactivity and have important implications for how neuropeptides shape defensive behaviors.

## Methods

### 1. Participants and Study Design

#### 1.1 Participants

Following evaluated recruitment criteria, 111 healthy volunteers (*n* = 37 per treatment group) were included in the final analysis (details see CONSORT flowchart in S1 Fig). Sample size was a priori determined using G*Power for a Treatment (3) × Sex (2) × IsThreaten (2) × Physical Stimulus Velocity (5 levels, from very slow to very fast; see Section 3.3 for detailed classification) mixed-design ANOVA, resulting in a required sample size of 78 (*f* = 0.20, *α* = 0.05, power = 0.95).

Eligible participants had normal or corrected-to-normal vision, no history of psychiatric or neurological disorders, and no current medication or substance use (see Supplementary Method S1 for full details). The study was approved by the local ethics committee of University of Electronic Science and Technology of China (UESTC), conducted according to the Declaration of Helsinki, and pre-registered (ClinicalTrials.gov ID: NCT06329063, NCT06329076). All participants provided written informed consent and received monetary compensation (140 RMB).

In a randomized, double-blind, placebo-controlled pharmacological behavioral and eye-tracking design, participants were randomly assigned to receive either AVP, LT, or PLC (mean age ± SD: 22.33 ± 2.22 years; no significant age differences between groups, *F*(2,108) = 2.13, *p* = .12).

### 2. Experimental Procedures

#### 2.1 Baseline Assessments

Prior to drug administration, participants completed validated Chinese versions of standardized questionnaires assessing affective states and fear-related traits, including the Positive and Negative Affect Schedule (PANAS) [35]; State-Trait Anxiety Inventory (STAI) [36]; Liebowitz Social Anxiety Scale (LSAS) [37]; and Animal Fear Questionnaire (AFQ) [38]. Blood pressure and heart rate were monitored at three time points: pre-administration, peak drug effect (45 min post-administration for AVP group, 90 min post-administration for LT group), and post-experiment (similar in [21,39], **Fig 1A**) to control for unspecific cardiovascular effects of the treatments.

**Fig 1.**
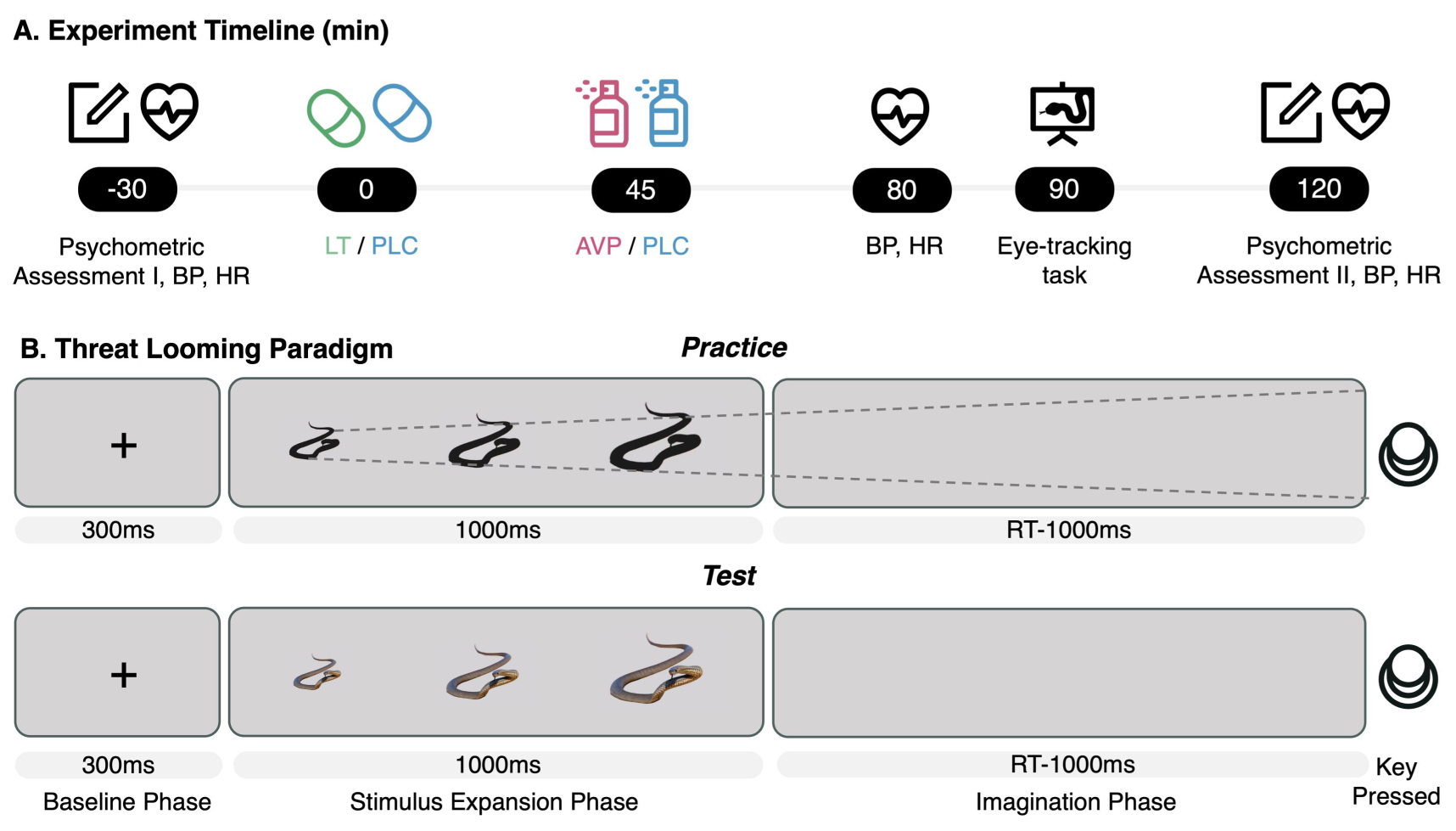
Experimental timeline and task structure. (A) Sequence of drug administration and behavioral testing. (B) Single trial structure of the threat looming task.

#### 2.2 Drug Administration

Treatment doses were selected based on previous research demonstrating efficacy in modulating emotional processing including fear-related processes: 20 IU AVP [39] and 50 mg LT [19–21].

We employed a novel spray-based oral delivery method for AVP [40] to prevent direct entry into the olfactory system and better isolate peripheral effects [41]. Following our validated protocol, participants received six alternating 0.1 ml puffs (total volume 0.6 ml, Supplementary Method S2). Placebo sprays were identical in composition but without AVP. Losartan (LT) and its placebo were administered in visually matched capsules containing either 50 mg Losartan or no active ingredient.

To ensure effects of both treatments during peak plasma windows (based on established pharmacokinetic profiles showing peak effects at 45 min for AVP and 90 min for LT) and maintain double-blinding during tasks, we implemented a two-stage administration protocol:

- Group 1: 50 mg LT capsule (90 min pre-task) + PLC spray (45 min pre-task)
- Group 2: PLC capsule (90 min pre-task) + PLC spray (45 min pre-task)
- Group 3: PLC capsule (90 min pre-task) + 20 IU AVP spray (45 min pre-task)

All treatments were administered by research staff blind to treatment conditions. Successful blinding was confirmed, as participants’ post-experiment treatment guesses did not exceed chance level (*χ*²(2) = 0.83 *p* = .36).

#### 2.3 Experimental Task

We employed a validated threat looming paradigm [13] using color photographs of threatening (snakes, spiders) and non-threatening (butterflies, rabbits) animals (40 images per category). Each trial (**Fig 1B**) began with a fixation cross that disappeared at 300 ms, followed by a looming stimulus that expanded at a constant velocity and disappeared at 1300 ms. After its disappearance, participants imagined its continued approach and pressed a key at their judged moment of collision. The judged Time-to-Collision (jTTC) was measured from stimulus onset (300 ms) to this key press. A confirmation beep was followed by a jittered inter-trial interval (1000-3000 ms). The initial stimulus size (20% or 30% of screen width) and its expansion velocity (determined by one of five actual Time-to-Collision (aTTC) values: 3.0, 3.5, 4.0, 4.5, and 5.0 s) were both varied independently across trials.

The experiment consisted of 160 trials across two blocks, comprising a full factorial design of aTTC (5 levels), stimulus category (4 levels), and initial size (2 levels). Participants completed practice trials using animal silhouettes prior to the main task. The task lasted approximately 35 minutes, and pupillary responses were recorded throughout.

### 3. Data Acquisition and Preprocessing

#### 3.1 Behavioral Data Preprocessing

Trial-level jTTC data were preprocessed in two stages. First, trials with technical artifacts or incomplete responses were excluded. Second, trials exceeding ±5 SD from each participant’s mean jTTC were classified as outliers and removed. This conservative criterion preserved potential treatment-induced variations while flagging extreme deviations, following methodological recommendations for pharmacological studies [42,43].

#### 3.2 Pupillary Data Preprocessing

Pupil size and movement from the right eye were recorded at 2000 Hz using an EyeLink 1000 Plus system (SR Research) with a display resolution of 1280 × 1024 pixels. Participants were positioned 57 cm from the monitor using a chin rest, and a 9-point calibration procedure was performed at the beginning of each experimental block to ensure optimal tracking accuracy.

Pupil size was recorded as pixel area and converted to diameter (*d*) using a validated formula *d* = *αLφ,* where *α* is an empirical scaling factor, *L* denotes the pupil-to-camera distance, and *φ* is the visual angle [44]. Data were preprocessed to remove signal loss, blink artifacts, physiologically implausible values, and statistical outliers beyond ± 5 SD from the trial mean, in accordance with established guidelines [28]. Finally, the cleaned pupillary signals were down-sampled into 10 ms bins to reduce high-frequency noise while preserving physiologically relevant fluctuations [45].

#### 3.3 Stimulus Dynamics

To optimize analysis, we computed a composite metric—Physical Stimulus Velocity (PSV)— integrating looming motion parameters into a single ecologically meaningful measure. Approach velocities (179.2-341.3 pixels/s) were categorized into five intervals: very slow (V1: 179.2-211.6), slow (V2: 211.6-244.0), medium (V3: 244.0-276.4), fast (V4: 276.4-308.8), and very fast (V5: 308.8-341.3 pixels/s, right-closed). These categories served as levels in subsequent analyses.

### 4 Analysis Framework

#### 4.1 Behavioral Analysis

To quantify the psychophysical characteristics of time perception, we employed a model-based approach: (1) a Linear Model (jTTC = *α* + *β* * aTTC), which served as a baseline assuming a linear relationship between subjective and objective time; (2) a Power Law Model (jTTC = *α* * (aTTC)^*β*), based on Stevens’ Power Law [46], which captures the canonical nonlinear relationship between perception and physical magnitude. In both models, *α* represents scaling parameters, while *β* reflects the rate of change (linear) or degree of nonlinearity (power law) in time perception.

For each participant and condition, model parameters (*α*, *β*) were estimated by iterative optimization, minimizing the sum of squared errors to obtain the best fit. The coefficient of determination (R²) was computed for each fit. To evaluate which model best accounted for the data across the dataset, we compared them using the Bayesian and Akaike Information Criteria (BIC and AIC) [47].

Finally, to examine the effects of Treatment, IsThreaten, and Sex on time perception parameters, we constructed Linear Mixed-Effects models using the estimated parameters (*α* and *β*) as dependent variables. Each model included fixed effects of Treatment, Sex, IsThreaten, and their possible two-way interactions. A random intercept for each participant was included to account for individual differences. The significance of the fixed effects was analyzed to determine whether the experimental manipulations modulated the temporal perception parameters.

#### 4.2 Pupil Analysis

##### (1) Baseline Pupil Analysis and Correction

Considering potential pharmacological effects on absolute pupil diameter, we first assessed baseline pupil diameter (0-300 ms) using a two-way ANOVA with Treatment and Sex as factors. All subsequent analyses were baseline-corrected by subtracting the mean baseline value from each timepoint to isolate task-evoked responses.

##### (2) Event-Locked Analysis

To compare mean pupillary responses across distinct phases of threat processing — from sensory encoding to decision preparation — we defined three temporal windows based on task structure and threat processing dynamics [32,48]:

(1). Stimulus Presentation (300-1300 ms; encoding of the approaching stimulus during active visual processing); (2). Early Imagination (first 500 ms post-offset; immediate transition to mental representation); (3). Late Imagination (last 500 ms pre-collision; decision-related preparatory processes).

A repeated-measures ANOVA was conducted with Treatment and Sex as between-subject factors, and Phase, IsThreaten, and Physical Stimulus Velocity (PSV) as within-subject factors. Trials shorter than 2300 ms (2%) were excluded to ensure complete phase coverage across all windows.

##### (3) Time-Normalized Dynamic Analysis

Variable trial durations — primarily due to individual differences in time-to-collision estimation — precluded direct comparison of pupillary dynamics across trials. Temporal normalization was therefore employed to align all trials to a common time scale. Following predefined quality criteria (Supplementary Methods S4), 9.65% of trials were excluded, with no differences in the percentage between treatment groups (*χ*²(2) = 0.46, *p* = .53). The remaining trials were processed using piecewise linear time warping to map the fixed stimulus epoch (300-1300 ms) to [0, 0.5] and the variable imagination epoch to [0.5, 1.0] on a unified scale, followed by smoothing of the normalized time series using cubic B-splines [49–51].

Functional linear mixed models (FLMM) were fitted [52] to examine the influence of Treatment, Sex, IsThreaten, and PSV on pupillary response over normalized time. The initial model followed a full factorial design:

Baseline-corrected Pupil Diameter ∼ Treatment * Sex * IsThreaten * PSV * Normalized_Time + (Normalized_Time + IsThreaten + PSV | Subject)

To ensure interpretability and avoid overparameterization, non-significant higher-order interactions were pruned using likelihood ratio tests [53], resulting in a final model that retained only effects up to three-way interactions.

Together, these analyses provide a comprehensive characterization of pupillary responses: baseline correction isolated pharmacological effects on tonic arousal, event-locked ANOVA identified phase-specific mean differences, and time-normalized FLMM captured full response trajectories across task progress. This multi-faceted approach distinguishes nonspecific drug effects, enables precise stage-specific inference, and reveals holistic temporal dynamics.

#### 4.3 Integrated Pattern Analysis

To identify treatment-specific psychophysiological phenotypes that reflect the integrated coordination of behavioral and pupillary systems during threat processing, we developed a comprehensive analytical framework combining Functional Principal Component Analysis (FPCA) [54], Clustering [55], and Hidden Markov Models (HMM) [56,57] (**Fig 2**). This approach captures both continuous temporal dynamics and discrete state transitions, aiming to reveal emergent, integrated phenotypes beyond simple correlations between systems. The analysis consisted of three sequential steps:

**Fig 2.**
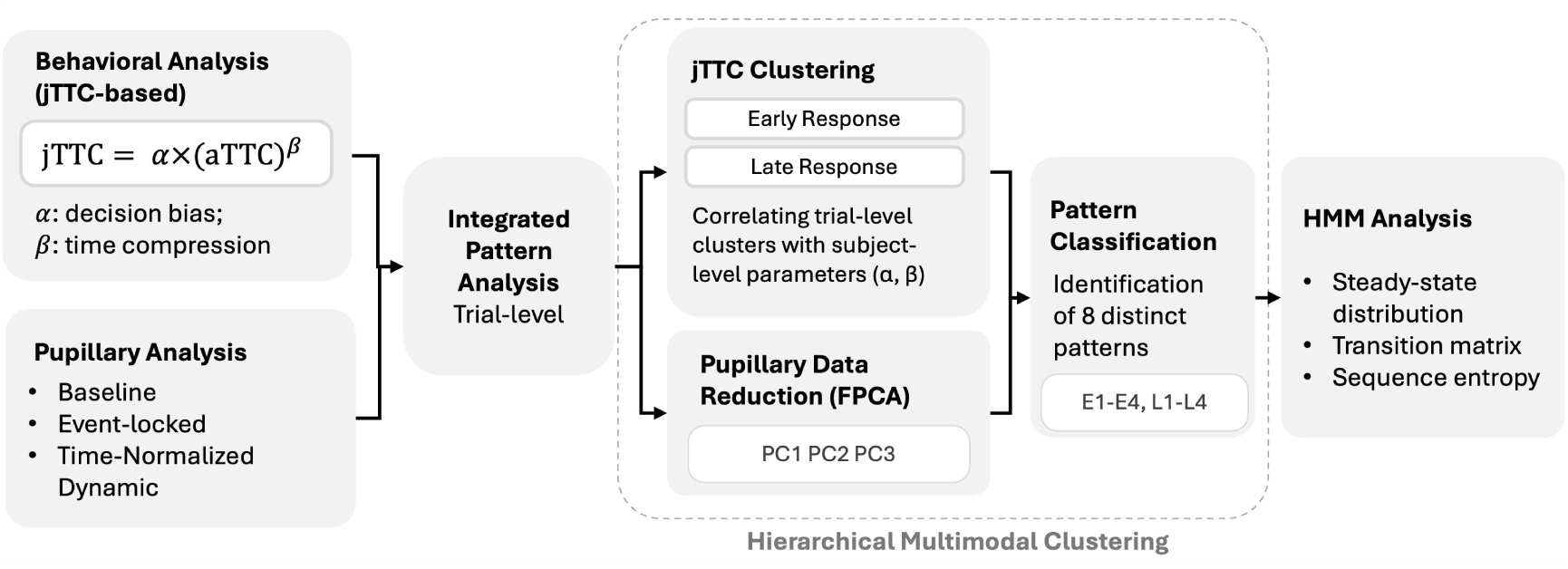
Analysis workflow. **First,** separate behavioral and pupillometric analyses were conducted. Behavioral responses were characterized using the power-law model jTTC = jTTC = *α* * (aTTC)^*β*. Pupillary analysis included baseline pupil size, event-locked responses, and time-normalized dynamics. **Second,** features were integrated through a hierarchical approach: trials were clustered based on jTTC values, with subgroup proportions correlated to subject-level parameters (*α*, *β*). FPCA reduced dimensionality of pupillary data, **followed by** Hidden Markov Modeling (HMM) to characterize state dynamics, including steady-state distributions, transition matrices, and sequence entropy.

##### (1) Functional Principal Component Analysis (FPCA)

Using FPCA, which better accounts for temporal continuity compared to traditional PCA [58], we extracted principal temporal response patterns (eigenfunctions) from time-normalized pupillary data within each trial. This dimensional reduction approach yielded multiple functional components per trial that collectively characterized the complete temporal dynamics of pupillary responses. We retained principal components that explained over 85% of total variance (individual components >5%) and examined their associations with experimental conditions, individual characteristics, and pupillary features during and after stimulus presentation.

##### (2) Hierarchical Clustering Integrated with jTTC and FPCA Components

To classify trials into distinct response patterns based on their behavioral-physiological characteristics, we integrated the FPCA components with behavioral features (jTTC) using a hierarchical k-means clustering framework. Given their different measurement scales, we first clustered on log-transformed jTTC, then on FPCA components within each behavioral subgroup. Optimal cluster numbers were determined using Calinski-Harabasz index and silhouette coefficients, with clustering stability verified through bootstrap resampling (*n* = 1000, stability index > 0.85) and cross-validation (accuracy > 0.80). To further determine whether these behavioral subgroups (trial-level) reflected stable computational traits (subject-level), we correlated the individual participants’ proportion of trials in each subgroup with their fitted power law parameters (*α* and *β*).

##### (3) Hidden Markov Modeling for Transition Dynamics

First-order Hidden Markov Models (HMMs) were applied to model transitions between response profiles across trials. Analysis encompassed three complementary aspects: First, the steady-state distribution was compared across experimental conditions using chi-square tests supplemented by standardized residual analysis; Second, transition probability matrices were estimated to capture state dynamics, with bootstrap methods (*n* = 1000) identifying significant transition patterns within each treatment group; Finally, the complexity of sequential pattern transitions was quantified through entropy measures applied to continuous three-state sequences across different experimental manipulations.

This integrated approach fuses continuous pupillary dynamics with behavioral data, identifying distinct, treatment-modulated response profiles and characterizing their temporal evolution during threat processing.

#### 4.4 Analytic Software and Correction

Behavioral analyses and event-locked phase analyses were conducted using SPSS 26. Time-normalized dynamic analyses and integrated pattern analyses were implemented in Python v3.12 using scientific computing libraries (pandas, numpy, scipy, statsmodels, scikit-learn) with custom implementations for functional data analysis. For all analyses, effects were considered significant at *p* < .05 with Greenhouse-Geisser correction for sphericity violations. Multiple comparisons were controlled for false discovery rate using the Benjamini-Hochberg method [59] (*α* = 0.05).

## Results

### 1 Subjective Anxiety and Potential Confounders

The three groups (LT, PLC, and AVP; *n* = 37 each) did not differ in age, mood, animal fear, or personality traits (all *p*s > .13, S1 Table). A 2 (Treatment: LT vs. PLC) × 2 (Sex: male vs. female) × 2 (Time: pre vs. post) mixed ANOVA with State Anxiety as the dependent variable revealed a significant Treatment × Time interaction (*F* = 4.11, *p* = .047, ^2^ = 0.055). Between-group comparison showed lower post-task state anxiety level in the LT group compared to PLC (*p* = .012, *d* = 0.51) which was mirrored in within-group analyses indicating significant anxiety reduction in the LT group (pre: 37.51 ± 1.10, post: 33.39 ± 1.36, *p* = .005, *d*z = 0.58) but not in the PLC group (pre: 38.44 ± 1.10, post: 38.36 ± 1.35, *p* = .95, *dz* = 0.02). This may suggest an anxiolytic effect of LT **(Fig 3A)**. No such pattern emerged for AVP (Treatment × Time, *p* = .78). Critically, the anxiety-reducing effect of LT did not differ by sex (*p*s > .41).

**Fig 3.**
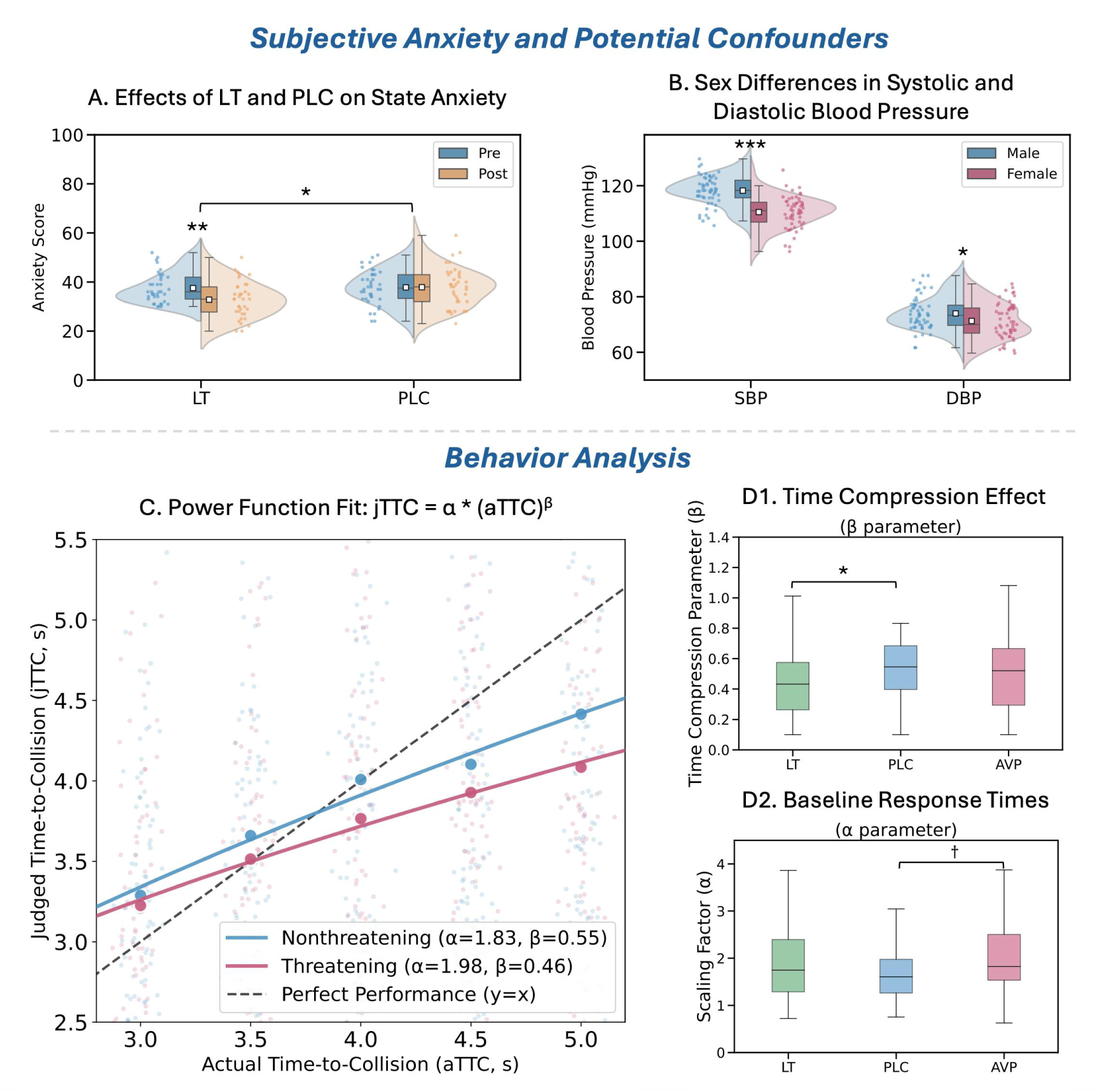
Subjective anxiety, potential confounders, and behavioral results. **(A) State anxiety scores pre- and post-experiment** in the LT and PLC groups. The LT group showed a significant reduction in anxiety at post-test. **(B) Systolic (SBP) and diastolic (DBP) blood pressure measurements**, revealing consistently higher values in males than in females. Both panels are presented as raincloud plots, where points represent mean scores per participant, the central line in the box indicates the median, and the box bounds represent the interquartile range. (C) Nonlinear fitting of aTTC and jTTC using the power-law model jTTC = *α* * (aTTC)^*β*. Participants generally underestimated time-to-collision, with accelerated underestimation as aTTC increased, particularly under threat conditions. Points represent mean jTTC per participant (threat vs. non-threat), jittered (5%) to avoid overlap. The dashed line y = x indicates accurate estimation. (D1) Parameter *β* (temporal scaling: *β* > 1 indicates expansion, *β* < 1 indicates compression). LT group exhibited significantly stronger time compression than PLC. (D2) Parameter *α* (general estimation bias: *α* > 1 indicates overestimation, *α* < 1 indicates underestimation). AVP group showed a tendency toward overestimation compared to PLC (*p*= .078). \**p* < .05,** *p* < .01, \*\*\**p* < .001, † marginal significance.

Cardiovascular parameters (systolic blood pressure, diastolic blood pressure, and heart rate) were monitored at three time points: baseline, drug peak effect (80 minutes post-LT; 35 minutes post-AVP), and post-task. Mixed ANOVAs revealed only a main effect of sex, with males showing higher resting blood pressure than females (SBP: 118.17 ± 0.74 vs. 110.34 ± 0.74 mmHg, *p* < .001; DBP: 73.66 ± 0.85 vs. 71.18 ± 0.85 mmHg, *p* = .041) **(Fig 3B).** No significant treatment effects or interactions were found at any time point (all *p*s > .10).

### 2 Time-to-collision Analysis

Model comparisons revealed that the power law model provided a superior fit to the linear model in most conditions (69.4%). The compression parameter *β* was consistently below 1 (range: 0.342-0.653, mean = 0.495), together with an average 1.55-fold increase in absolute perceptual error as aTTC increased from 3s to 5s, indicating systematic time compression that accelerated with longer aTTCs. Linear mixed-effects (LME) analyses revealed a significant reduction in *β* (indicating enhanced time compression) for threatening compared to nonthreatening stimuli ((*β* = -0.091, *p* = .06; **Fig 3C**), under LT compared to PLC (*β* = -0.149, *p* = .033; **Fig 3D1**), and in females compared to males (*β* = -0.167, *p* = .016). No significant reduction in *β* was observed for AVP compared to PLC (*β* = -0.045, *p* = .52).

The scaling parameter *α* (where *α* = 1 indicates accurate estimation) showed substantial individual differences (range: 0.857-2.341, mean = 1.534). LME analyses revealed a trend toward increased *α* (indicating a tendency for time overestimation) under AVP compared to PLC (*β* = 0.504, *p* = .078; **Fig 3D2**). No other effects reached significance.

### 3 Pupil Analysis

#### 3.1 Baseline Analysis

Analysis of pre-stimulus baseline pupil diameter revealed a significant main effect of treatment (*F* = 4.31, *p* = .016), with the AVP showing larger diameters than the PLC group (*p* = .011), the LT did not significantly differ from either group (*p*s > .104). No sex differences were observed (*p*s > .2).

This pharmacological effect persisted throughout the entire experiment, as confirmed by analysis of uncorrected data across trials (*F* = 3.98, *p* = .023; **Fig 4A**). After baseline correction, these general differences were eliminated (*p*s > 0.12) allowing examination of task-specific pupillary changes.

**Fig 4.**
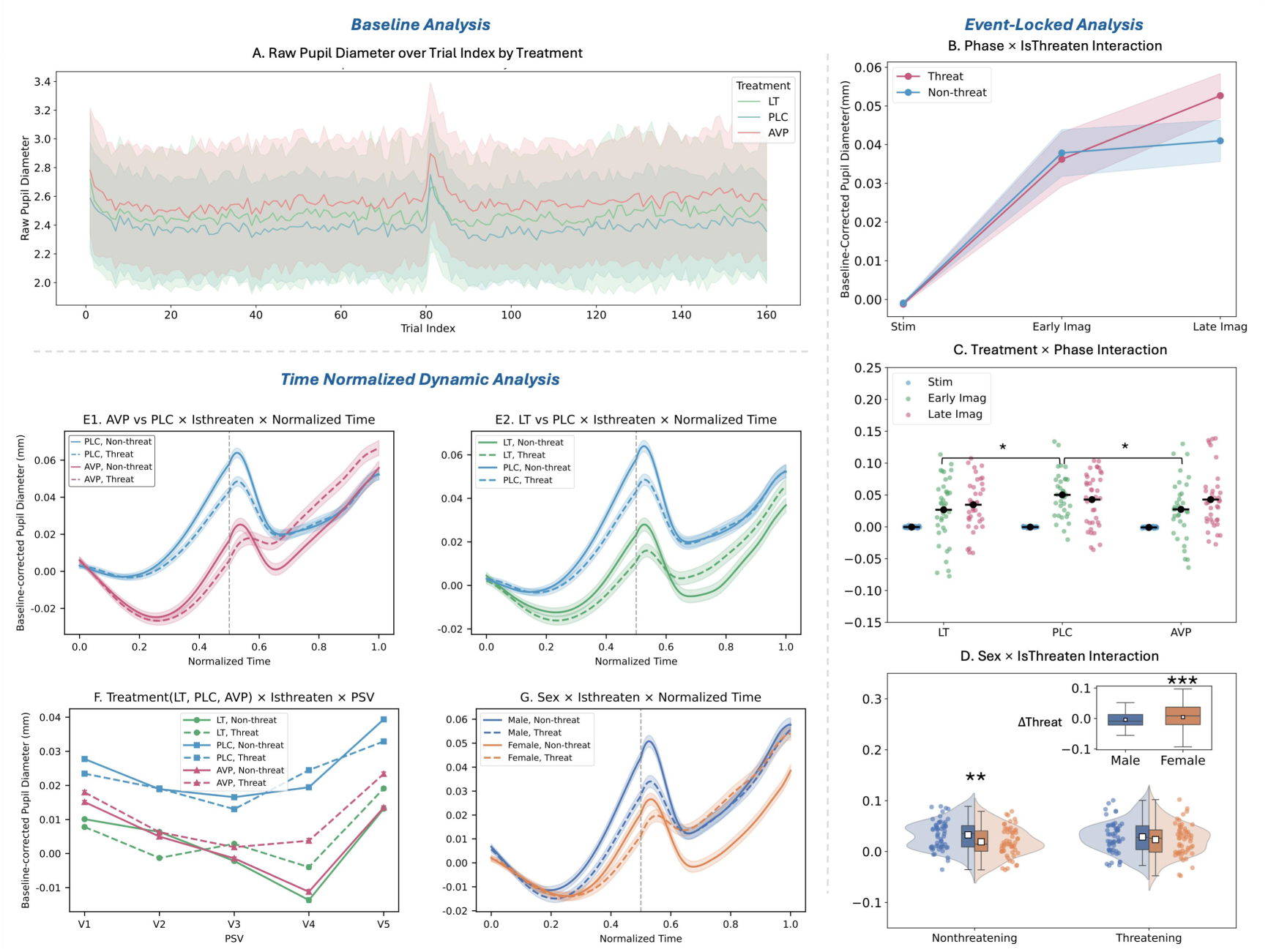
Pupillary Measurement: Baseline (A), Phase-Locked (B-D) and Normalized Time Dynamic Analyses (E-G). **(A)Baseline pupil size:** The AVP group had a significantly larger uncorrected pupil diameter than the PLC group. The LT group did not differ from either. **(B) Phase × Isthreaten:** Threat evoked a sustained dilation, while non-threat elicited a phasic dilation followed by a plateau. Responses to threat significantly exceeded non-threat in the late imagination phase. **(C) Phase × Treatment:** The PLC group response was significantly larger than both AVP and LT during the early imagination phase. The AVP group exhibited sustained dilation across phases, while PLC and LT responses plateaued from early to late imagination. **(D) Sex × Isthreaten:** Males had significantly larger pupils than females specifically during the non-threat condition (no sex difference under threat). The inset shows a significantly greater threat-specific response (threat - non-threat) in females. **(E1) AVP vs. PLC × Isthreaten × Time:** PLC > AVP during stimulus approach; this pattern reversed to AVP > PLC after stimulus offset under threat. **(E2) LT vs. PLC × Isthreaten × Time:** PLC > LT across time. After offset, threat > non-threat for LT, while PLC showed no threat-non-threat difference. **(F) Treatment × Isthreaten × PSV:** Pupillary velocity showed a V-shaped profile. The nadir occurred earlier for PLC (V3) than for drug groups (V4). AVP showed threat > non-threat from V1-V5, LT from V3-V5, while PLC showed no differential sensitivity. **(G) Sex × Isthreaten × Time:** Male > female during stimulus approach; this difference disappeared after stimulus offset under threat. **\* *p* < .05, ** *p* < .01, \*\*\**p* < .001.**

#### 3.2 Event-locked Analysis

Analysis of baseline-corrected responses revealed significant main effects of Phase (*F* = 41.97, *p* < .001) and PSV (*F* = 19.92, *p* < .001). Pupils dilated from stimulus presentation to early imagination (Present vs Early: *p* < .001) and stabilized during late imagination (Early vs Late: *p* = .22). Extreme velocities (V1/V5) elicited larger responses than moderate velocities (V2-V4, all *p*s < .01).

A Phase × IsThreaten interaction (*F* = 16.20, *p* < .001; **Fig 4B**) revealed that threatening stimuli induced progressive dilation across phases (Stim < Early < Late, *p*s < .05), while non-threatening stimuli showed initial dilation followed by plateau (Stim < Early = Late, *p*s < .01). During late imagination, threat-induced pupil dilation exceeded non-threatening responses (*p* = .004).

A Phase × Treatment interaction (*F* = 2.93, *p* = .022; **Fig 4C**) showed that during early imagination, both LT and AVP reduced responses as compared to PLC (*p*s < .05), while no group differences were observed during stimulus presentation or late imagination (*p*s > .22). Across phases, AVP group showed progressive dilation (Stim < Early Imag < Late Imag, *ps* < .05), LT and PLC groups showed initial dilation followed by plateau (Stim < Early Imag ≈ Late Imag, *p*s < .01).

The IsThreaten × Sex interaction (*F* = 18.513, *p* < .001; **Fig 4D**) revealed that males showed larger responses than females to non-threatening stimuli (*p* = .003), but no sex differences under threat (*p* = .84). Females exhibited enhanced responses to threatening versus non-threatening stimuli (*p* < .001), while males’ responses remained consistent across both conditions (*p* = .10).

#### 3.3 Time-Normalized Dynamic Analysis

Extending event-locked findings, functional linear mixed models on baseline-corrected pupil diameter across normalized time revealed more nuanced patterns.

Overall pupil dilation increased with normalized time (*β*=0.010, *p*=.030) and threat-induced constriction (*β*=-0.016, *p*< .001). PSV demonstrated a V-shaped velocity-dependent pattern, with maximal dilation at extreme velocities (V1/V5: *β* = 0.016/0.018, *p*s<.001), moderate dilation at V2 (*β* = 0.005, p<.001), and no change at V4 relative to V3 (*p* = .63).

Treatment × IsThreaten × Normalized Time interaction (*β*s = 0.015-0.024, *p*s < .05; **Fig 4E1,E2**) showed that during stimulus approach (t=0-0.5), all groups exhibited threat-induced constriction (*p*s<.01), but after stimulus offset (t=0.5-1), threat processing diverged: PLC showed no threat/non-threat difference while both LT and AVP maintained enhanced dilation under threat versus nonthreat (*p*s < .001). Notably, between-group comparisons revealed that the LT group maintained overall smaller response than PLC, whereas pupil response was smaller in the AVP compared to the PLC group during stimulus presentation but showed enhanced dilation post-stimulus specifically for threat (*β* = 0.008, *p* < .001).

A Treatment × IsThreaten × PSV interaction (**Fig 4F**) indicated that the PLC group showed the largest overall responses compared to both AVP and LT groups (*β* s = 0.017-0.041, *p*s < .05). AVP group showed larger responses to threatening versus non-threatening stimuli across all velocities (V1-V5: *β*s = 0.0028-0.0099, *p*s < .001), and LT selectively increased threat responses only at faster velocities (V3-V5: *β* = 0.005-0.0097, *p*s < .001). PLC group showed no velocity-specific threat sensitivity. All groups maintained the V-shaped velocity effect, with drug groups reaching their lowest response at V4 while the PLC group reached its lowest response at V3.

A significant Sex × IsThreaten × Normalized Time interaction (*β* = 0.025, *p* < .001; **Fig 4G**) emerged. During stimulus approach, males showed larger pupil response than females across both threat and nonthreat conditions (*p*s < .001). After stimulus offset, however, the sex difference persisted only for non-threatening stimuli (*β* = 0.0221, *p* < .001) and was abolished under threat (*p*=.27). Crucially, females exhibited greater dilation to threatening versus non-threatening stimuli post-stimulus (*β* = -0.0193, *p* < .001), while males showed no threat-related differentiation (*p*=.31).

### 4 Integrated Behavioral-Pupillary Response Profiles

Eight distinct response profiles were identified through a hierarchical clustering procedure. First, clustering trials based on log jTTC revealed two behavioral subgroups: an early-response group (E: 3.12 ± 0.57 s) and a late-response group (L: 5.80 ± 1.98 s). Correlations with power law parameters confirmed associations with distinct subject-level traits in time estimation: the early-response proportion negatively correlated with both *α* (*r* = −0.45, *p* < .001) and *β* (*r* = −0.36, *p* < .001), indicating systematic underestimation and stronger time compression; conversely, the late-response proportion was positively correlated with both *α* (*r* = 0.45, *p* < .001) and *β* (*r* = 0.36, *p* < .001), indicating the opposite pattern of overestimation and weaker compression.

Second, FPCA decomposed pupillary dynamics into three core components (PC1-3, **Fig 5A**) accounting for 87.51% of total variance, which were differentially modulated by experimental conditions and individual traits (summarized in **Table 1**; full analysis in Supplementary Results: Functional Principal Components, S2-S3 Fig). Based on the full analyses the components were interpreted as PC1: Sustained Vigilance, PC2: Proactive Engagement, reflecting active preparation for threat, and PC3: Cognitive Shift, capturing the transition from perception to imagination.

**Fig 5.**
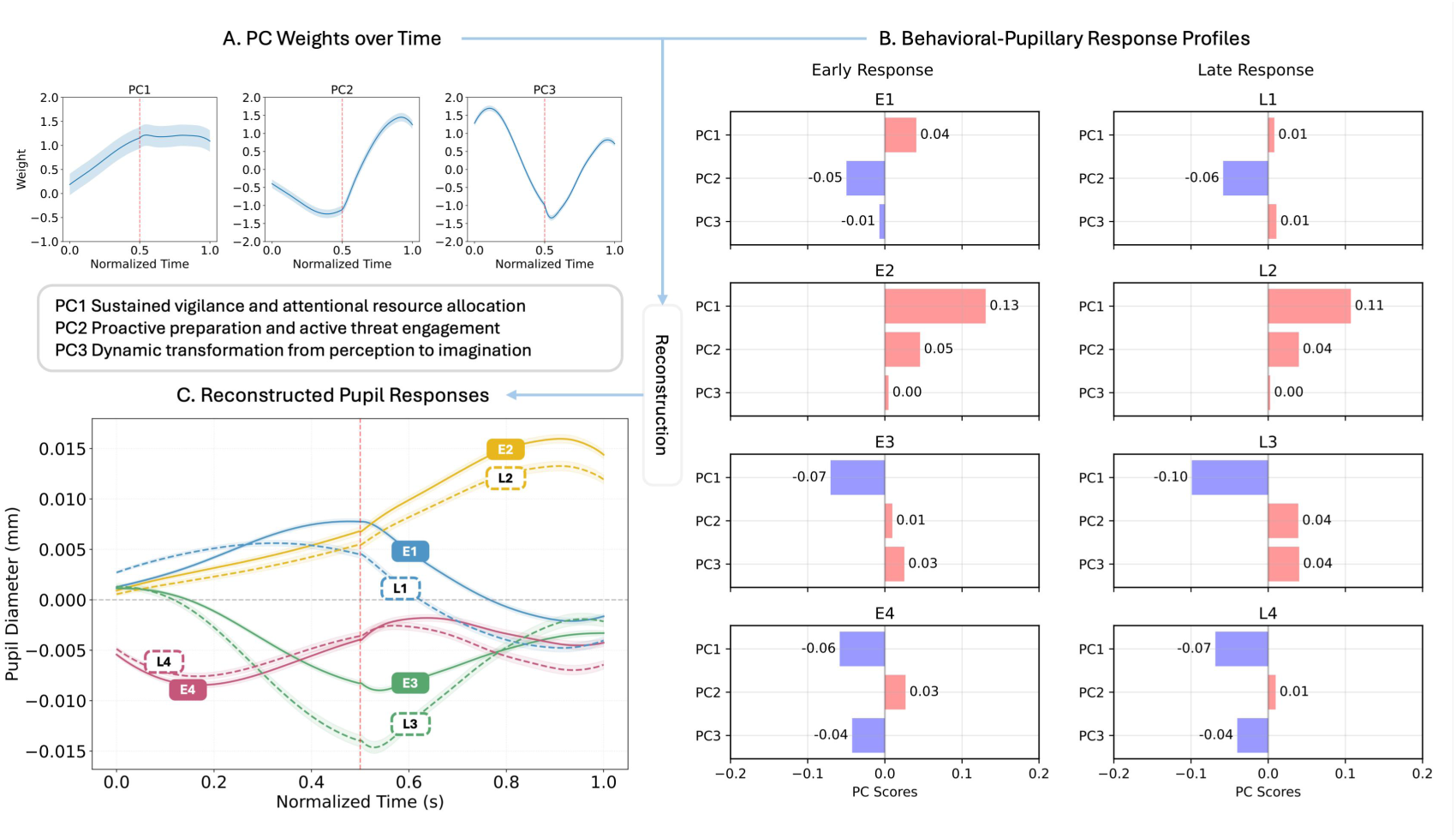
Decomposition and profiling of integrated behavioral-pupillary responses. **(A) Three dominant functional principal components (PCs) underlying pupillary dynamics.** Eigenfunctions *φ*(t) for each component are displayed, with their sign and magnitude defining the functional interpretation: PC1 (Sustained Vigilance) shows sustained positive weighting; PC2 (Proactive Engagement) transitions from negative to positive weighting; PC3 (Cognitive Shift) exhibits a multiphasic (positive-negative-positive) pattern. Individual trial responses are reconstructed as a linear combination of these components: Pupil Diameter(t) ≈ Grand Mean(t) + (Score₁× *φ₁*(t)) + (Score₂ × *φ₂*(t)) + (Score₃ × *φ₃*(t)). **(B) Eight distinct response profiles from hierarchical clustering.** Profiles were identified by combining behavioral response subgroups (Early: E1–E4; Late: L1–L4) with clustering of PC. **(C) Reconstructed pupillary responses for each profile.** Curves illustrate the characteristic pupillary signature of each profile over time, reconstructed by combining the shared temporal eigenfunctions from (A) with the profile-specific component scores from (B).

**Table 1.** summarizes the characteristic features and experimental correlations of each component, along with their proposed functional significance.

| Principle Component | PC1 | PC2 | PC3 |
| --- | --- | --- | --- |
| Variation Explanation | 62.67% | 17.56% | 7.28% |
| Weight Time Function Features | Early steep rise followed by plateau | Initial inhibition, later strong activation | Early peak, mid-period trough, late recovery |
| Main Related Pupil Features | Stimulus-Evoked Response<br>(Pupil size during stimulus: $r=.615$, speed of dilating: $r=.65$) | Post-Stimulus Sustenance (Pupil size after stimulus: $r=.668$) vs. Stimulus Suppression (Pupil size during stimulus: $r=-.35$) | Rate Anticorrelation (Slower dilating during stimulus: $r=-.58$, Faster dilating after: $r=.34$) |
| Experimental Conditions | Female only:<br>Nonthreatening:<br>PLC>LT, PLC>AVP<br>Threatening:<br>PLC=AVP>LT<br>Velocities: V1,V5>V2-V4<br>(No male differences) | AVP>LT, AVP>PLC<br>Threat>Nonthreat<br>V1,V2<V3-V5 | Threat>Nonthreat<br>V1,V5<V2-V3 |
| Individual Differences | not significant | Low anxiety > High Anxiety | Early response < Late response |
| Possible Functional Significance | Sustained Vigilance and Attentional Resource Allocation | Proactive Preparation and Active Threat Engagement | Dynamic Transformation from Perception to Imagination |

Integrating behavioral (E/L) and pupillary (PC1-3 scores) features, a second-level clustering yielded eight stable profiles (E1-E4, L1-L4, **Fig 5B**), verified by bootstrap (stability index: 0.753 ± 0.125) and cross-validation (0.651 ± 0.114; S4 Fig). These profiles reflect distinguishable threat processing strategies (Table 2):

**Table 2.**
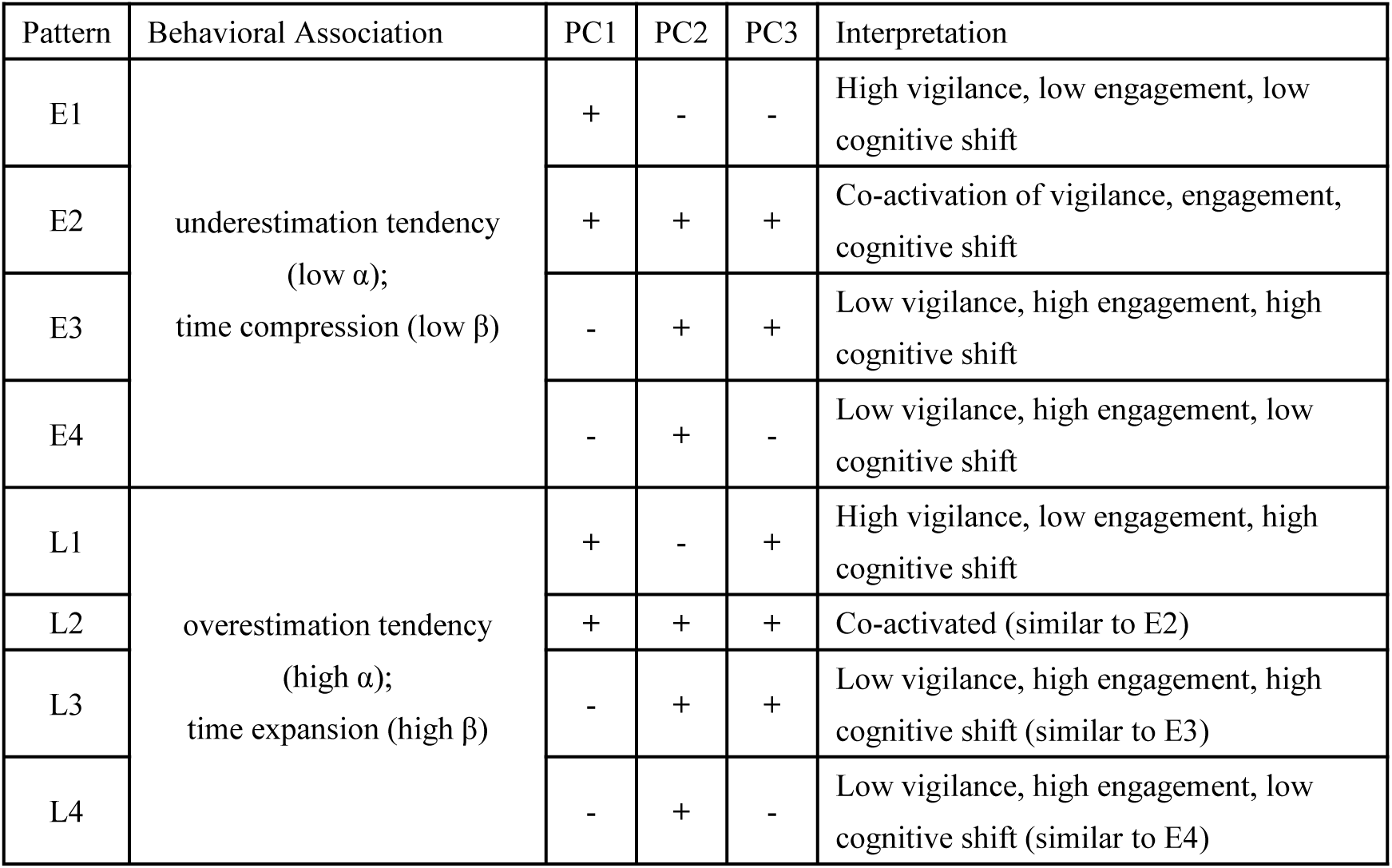
Characteristics of the eight behavioral-physiological profiles.

For instance, profile E1 was characterized by dominant sustained vigilance (high PC1) with suppressed proactive engagement and cognitive shift (low PC2/PC3). Profile E2 was characterized by co-activated vigilance, engagement, and cognitive shift (high PC1/PC2/PC3). Notably, late-response profiles L2-L4 shared the same directional patterns of PC scores as E2-E4 but differed in response magnitude, as shown in their reconstructed responses (**Fig 5C**).

#### 4.3 HMM Analysis

##### (1) Steady State Distribution

Residual analysis revealed a significant main effect of treatment (*χ*²(14) = 333.17, *p* < .001; **Fig 6A**). **Profile E1** (characterized by approach-phase dilation and post-stimulus constriction) was a dominant state that showing a significantly higher probability in PLC (*z* = 8.75) and a lower probability in AVP (*z* = -9.85). **Profile L3** (enhanced constriction during approach phase followed by attenuated constriction post-stimulus) defined the pharmacological interventions, exhibiting significantly higher probability in both AVP *(z* = 4.96) and LT (*z* = 3.53) groups compared to PLC (*z* = -8.32). **Profile L2** (sustained dilation) further distinguished the two active treatments, being higher in AVP (*z* = 3.57) and lower in LT (*z* = -2.69) **(see Fig 5 for each profile’s visual breakdown)**.

**Fig 6.**
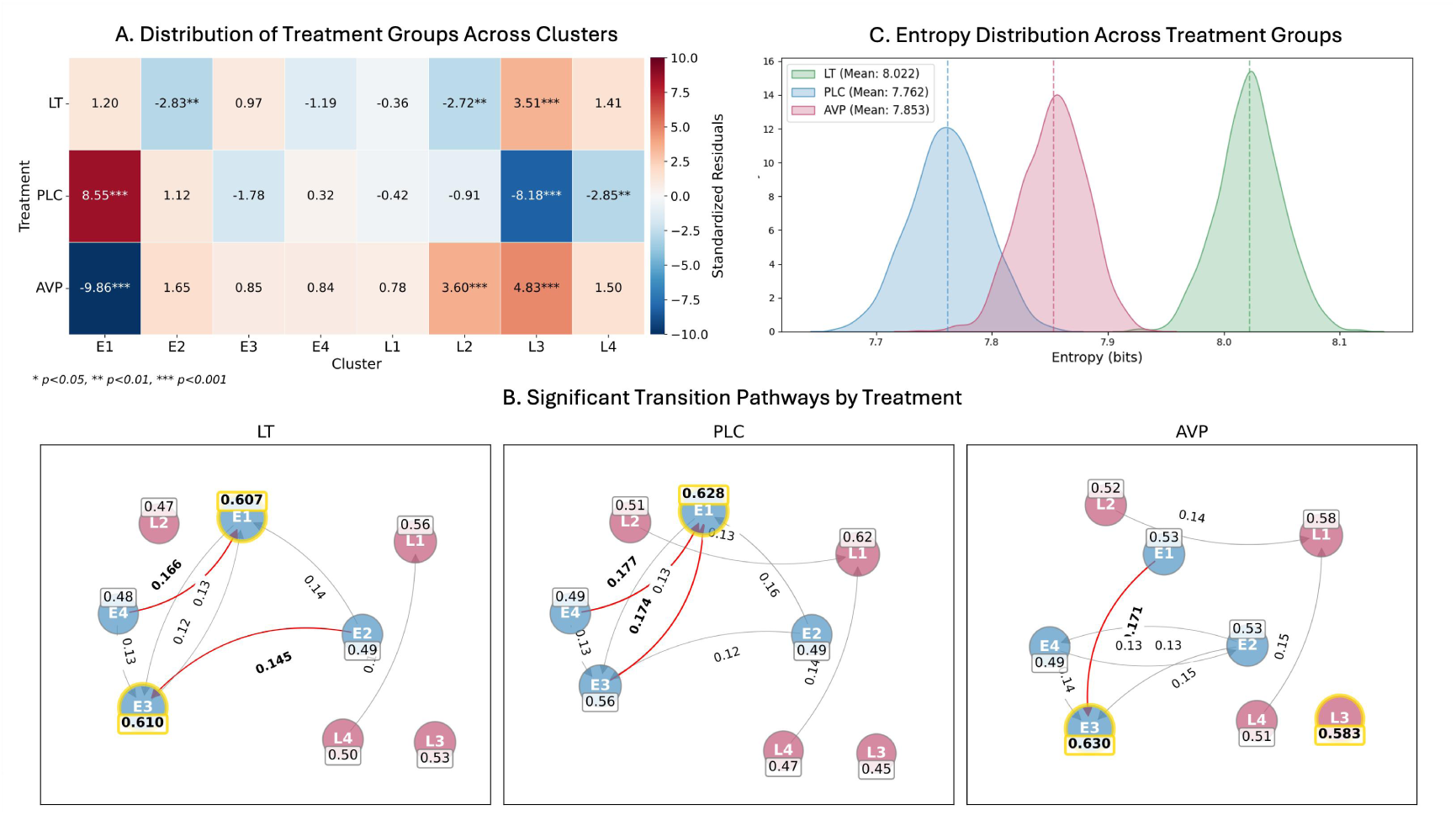
Hidden Markov Model (HMM) analysis of response patterns. **(A) Stationary distribution revealed treatment-specific deviations from expected state probabilities.** Profile E1 was significantly elevated in PLC and reduced in AVP. Profile L2 was increased in AVP and decreased in LT. Profile L3 was elevated in both LT and AVP but suppressed in PLC. Profile L4 was significantly lower in PLC (* *p* < .05, ** *p* < .01, *** *p* < .001). (B) Significant within-group transition pathways (permutation test, *p* < .05). Red arrows and bolded probabilities indicate significant transitions. Grey arrows denote non-significant transitions. Node labels show self-transition probabilities. Integration with between-group comparative analyses revealed the following key treatment effects on transition dynamics: **PLC** reinforced Profile E1 (sustained vigilance) as a dominant attractor state, through high self-persistence and convergent inflows from other states. **AVP** disrupted transitions toward E1 while facilitating transitions toward states of full component co-activation (e.g., L2) or threat-specific engagement (e.g., E2E3). **LT** selectively inhibited the emergence of the co-activated L2 state, as evidenced by a lack of significant convergent transitions compared to other treatments. **(C) Sequence entropy** differed significantly across treatments (LT > AVP > PLC), indicating that PLC produced the most predictable and stable sequences, whereas LT sequences were the most dynamic and unpredictable.

The sex effect (*χ* ² (7) = 250.84, *p* < .001; S6 Fig) showed males demonstrated elevated engagement in late-response profiles (L1 – L4; *z* = 1.81 – 6.86) while females showed reduced engagement (*z* = -1.92 to -7.27). Specifically, profile L1 was higher in males (*z* = 6.86) and lower in females (*z* = -7.27), while profile E3 was higher in females (*z* = 7.60) and lower in males (*z* = -7.17). The Isthreaten effect (*χ* ² (7) = 65.59, *p* < .001) revealed profile E1 exhibited lower engagement under threat (*z* = -3.17) than non-threat (*z* = 3.11), and profile E2 exhibited higher engagement under threat (z = 4.43) than non-threat (*z* = -4.35).

##### (2) Dynamics Transition

Analysis revealed treatment-specific remodeling of state transition pathways, which occurred despite consistently high self-transition probabilities (0.45–0.63) across all conditions (see S7 Fig for full statistics).

Specifically, the PLC group showed significantly increased probabilities in transitions towards E1 (e.g., E3→E1: *z* = 4.65; E4→E1: *z* = 2.49; all *p*s < .05). Conversely, the AVP group significantly suppressed transitions toward E1 (e.g., E1→E1: *z* = -4.30; E4→E1: *z* = -3.40) while promoting transitions involving late-response profiles (e.g., E4→L2: *z* = 2.92; L3→L3: *z* = 2.83). The LT group specifically exhibited reduced transitions toward the L2 profile (e.g., E2→L2: *z* = -2.32; E4 → L2: *z* = -1.99). Within-group transitions **(Fig 6B)** further supported these between-group findings and provided a more detailed architecture of significant pathways within each treatment (see Supplementary S2-S3 Table for full details, 95% CIs, and pairwise comparisons).

Sex comparisons (*χ*² = 250.85, *p*-adj < .001) revealed males exhibited increased transitions toward L1 (E1 → L1: *z* = 2.75; E3 → L1: *z* = 2.33) while females showed elevated transitions toward E3 (E1 → E3: *z* = 2.32; L4 → E3: *z* = 3.18). A significant threat modulation was also identified, enhancing the E3 → E2 transition under threatening conditions (*χ*² = 14.95, *p*-adj = .007).

##### (3) Sequence Entropy and Stability

Analysis of sequential patterns (**Fig 6C**) provided further evidence for treatment-induced destabilization of cognitive states, extending beyond transition probabilities to higher-order sequence inconsistency.

PLC group demonstrated the most predictable and stable sequences, exhibiting the lowest Markov entropy (M = 7.76 bits, 95% CI [7.70, 7.82]), which consistent with its tendency to stabilize around repetitive, E1-centric sequences (e.g., E1→E1→E1, *z* = 5.13, *p* < .001).

Conversely, both pharmacological treatments significantly increased sequence entropy (i.e., made sequences more random and less predictable), but did so through distinct mechanisms:

LT group produced the highest entropy (M = 8.02 bits, 95% CI [7.96, 8.08]), significantly exceeding both AVP and PLC (both *p*-adj < .001), indicating a generalized state of maximal inconsistency without promoting any specific alternative sequences.

AVP group exhibited intermediate entropy (M = 7.85 bits, 95% CI [7.80, 7.91]), significantly higher than PLC (*p*-adj < .001) but lower than LT (*p*-adj < .001). This reflects a partial disruption of the stable PLC dynamics, coinciding with its shift toward sustained E3 and L3 patterns.

Sex comparisons revealed significantly lower sequence entropy in females compared to males (diff = -0.05 bits, *t* = 42.55.13, *p* < .001); Threatening stimuli induced a subtle but significant reduction in sequence entropy compared to non-threatening conditions (diff = -0.02 bits, *t* = 20.13, *p* < .001), suggesting increased predictability of state sequences under threat. (Detailed analysis of specific 3-state sequences is provided in Supplementary Table S4.)

## Discussion

Our study investigated the behavioral and pupillary dynamics underlying the processing of looming threats — both during sensory approach and internal imagination — and how these processes are modulated by the neuropeptides vasopressin (AVP) and angiotensin II (LT). Using a randomized, placebo-controlled design integrating behavior and eye-tracking, we reveal that looming threat evaluation unfolds through distinct temporal stages and is modulated by neuropeptide signaling, which regulate processes of sustained vigilance and adaptive flexibility.

Our findings highlight three aspects of looming threat evaluation and its modulation:

(1) threat-induced time compression: we confirmed that the collision time for looming stimuli is underestimated (see also [13]) and our computational modeling revealed that the underestimation of collision time for threating stimuli (vs. nonthreat) is based on an accelerated rate of temporal compression (*β*), rather than a threat-induced modulation of constant bias (*α*). This threat-induced time-compression was further modulation by sex.
(2) dynamic pupillary responses during approaching threat and occlusion: a dynamic biphasic pupil response pattern characterized by sustained constriction during the visual approach (reaching minimum at stimulus disappearance), transitioning towards a release of constriction to progressive dilation prior to the expected (imagined) collision, which may reflect a dynamic interplay between vigilance and defensive responses.
(3) neuropeptide-specific modulation: AVP and LT exerted opposing on neural-behavioral states. AVP prolonged time perception by stabilizing hyper-vigilant states (e.g., L2) and increasing transitions into these states. LT, in contrast, promoted time compression and flexibility by suppressing transitions into L2 and increasing state-sequence entropy, suggesting a calmer processing mode.

### Threat-Induced Time Compression of Looming Threat

Building on and confirming earlier findings demonstrating that threat induces an underestimation of collision time [13], our computational modeling further revealed that this effect arises from an acceleration in the rate of temporal compression (*β*) — particularly for longer aTTCs — without changes in the overall constant bias (*α*). This indicates that threat modulates the gain of the estimation process rather than its starting point [60,61]. This pattern may reflect a selective increase in the pace of evidence accumulation to support expedited action, which align with existing theoretical frameworks proposing that motivational states influence decision-making model parameters [62,63], and indicate that threat enhances processing efficiency (drift rate) rather than lowering response caution (decision criterion) during looming.

Furthermore, pupillary responses revealed distinct temporal dynamics and threat-sensitive modulation. During the visual approach phase, threat elicited a sustained constriction that peaked at stimulus disappearance. This progressive constriction may reflect increasingly active content decoding, contributed to by a cognitively driven near reflex as participants attempted to foveate and resolve the details of the looming stimulus. However, the sustained, time-locked nature of the constriction suggests it this response may additionally reflect an integrated engagement of cognitive and oculomotor systems when evaluating looming stimuli [64,65]. Following stimulus offset, pupils transitioned to a progressive dilation, potentially reflecting a shift from external sensory processing to internal simulation. This dilation may suggest increasing autonomic arousal and anticipatory defense activation as the imagined collision nears [66,67] and active maintenance and estimation of the stimulus and its trajectory in working memory.

Critically, approach velocity significantly modulated these pupillary responses, exhibiting a V-shaped pattern across speeds. Maximum constriction occurred at intermediate velocities (V2 for threat, V4 for non-threat), while both the slowest (V1) and fastest (V5) velocities elicited greater dilation. We hypothesize that this pattern reflects a shift in defensive strategies based on spatial imminence [68]: detailed sensory analysis is prioritized at intermediate speeds via constriction [28], whereas extreme velocities associate with a shift in arousal under conditions of either low (V1) or high perceived relevance (V5) [69,70]. These findings underscore a continuous interaction between looming and semantic content in shaping visual and threat responses, aligning with broader models on changes in threat processing along the imminence spectrum [34,68].

The semantic content-related processing differences were further supported by results from the pattern analysis such that nonthreatening stimuli preferentially engaged Profile E1 whereas threatening stimuli robustly recruited E2, as evidenced by: (1) a higher-than-expected steady-state distribution in their respective profiles, (2) distinct patterns in the transition matrices, and (3) lower sequential entropy for the E1-dominated non-threat stimuli, indicating a more stable, deterministic state. Interpreting these profiles via principal component loadings **(Fig 5A-B**; **Table 2)** underscores a threat-related dual processing mode:

**Non-threatening Profile E1** (+, -, -) reflects an efficient surveillance mode characterized by sustained vigilance (PC1+) with negative engagement in proactive threat preparation (PC2-) or cognitive state transformation (PC3-). The associated pupillary response **(Fig 5C)** — initial expansion followed by contraction—is consistent with online monitoring and rapid disengagement after classifying a stimulus as non-critical, reflecting an adaptive resource conservation strategy [71].

**Threat-related Profile E2** (+, +, +) constitutes a high-alert threat mode involving the co-activation of vigilance (PC1+), proactive threat preparation (PC2+), and sustained internal simulation (PC3+). Its persistent pupillary dilation **(Fig 5C)** reflects ongoing resource allocation to threat anticipation, consistent with a state of heightened defensive readiness that facilitates exaggerated threat simulation and perceptual preparation [72].

Critically, both profiles are associated with early responses (short jTTC) and correlate negatively with *α* and *β* parameters, indicating that while E1 and E2 represent distinct computational pathways, they both facilitate a common adaptive output to looming stimuli: a behavioral tendency for underestimation of time-to-collision, which is further amplified in the case of threatening stimuli. This bridges our trial- and group-level findings: a general time compression (E1-driven) is significantly enhanced by threat (E2-driven).

### Sex Differences in Threat Processing Strategies

Sex modulated the described threat processing architecture, such that females exhibited a steeper rate of temporal compression (*β*: female < male), suggesting that women may adopt a more precautionary decision-making policy, favoring accelerated defensive responses under threat [73,74].

Pupillometric analyses suggested that while males demonstrated generally larger pupil diameters, only females displayed a significant threat-induced differentiation (threat > non-threat) following stimulus offset. This post-stimulus divergence in females may suggest enhanced engagement of internal, defensive simulation processes for threatening content [69]. Sustained pupil dilation – as observed in our experiment – has been associated with emotional arousal and increased cognitive load during such internal representations [75].

The sex-differential processing of threat is further underscored by findings from the pattern analysis (details see Supplementary Results: Sex Differences in Patterns and Fig 5A-C) which suggest a **male processing strategy (Profile L1)** reflected by state of broad, shallow monitoring that maintains readiness at the cost of detailed threat evaluation, resulting in delayed responses. In contrast, the **female-biased strategy (Profile E3)** may facilitate targeted internal modeling of threats after stimulus offset, supporting superior threat discrimination and promoting earlier, precautionary responses.

### Neuropeptide Modulation of the Looming Response

#### The Placebo Baseline: A Resource-Conserving Mode for Fading Threat Cues

Under placebo, the pupillary response to threatening stimuli was diminished relative to non-threatening ones during approach, and this discrimination vanished after offset **(Fig 4E1-E2)**. The integrated pattern was dominated by **Profile E1** (+, -, -), as evidenced by its high steady-state probability **(Fig 6A)**, a bias for transitions into E1 from other states **(Fig 6B)**, and the lowest sequential entropy (indicating high stability) **(Fig 6C)**. This profile is characterized by sustained vigilance (PC1+) with low investment in proactive threat preparation (PC2-) or extended internal simulation (PC3-), consistent with the reconstructed pupillary response of initial expansion followed by pronounced contraction **(Fig 5A, PC weight functions; B, profile characteristics; C, reconstructed pupil response)**.

#### Vasopressin Promotes a Hyper-Driven, Enhanced Simulation Mode

Vasopressin (AVP) administration reconfigured threat processing towards a state of higher engagement. Behaviorally, this manifested as a significant time-to-collision overestimation (*α* > PLC). Pupillometric analyses revealed that AVP specifically enhanced and sustained threat-discriminative responses during the imagery phase. This pattern was not modulated by variations in velocity suggesting an internal process that was maintained across external variations. Although the peripheral administration of a peptide like AVP—with its debated blood-brain barrier permeability — raises questions about central bioavailability, the significant behavioral and pupillary effects observed here align with a growing body of evidence indicating that peripherally administered neuropeptides can exert central actions through direct as well as indirect mechanisms, such as vagal nerve activation or engagement of receptor-mediated transport systems (details see also [76–78]).

Hidden Markov modeling (HMM) further identified that AVP suppressed the baseline E1 mode while promoting stability in a high-arousal state (**Profile L2** +, +, +) **(Fig 6A-B)**. Critically, L2 represents a co-activation of sustained vigilance and resource allocation (PC1+), proactive preparation for threat engagement (PC2+), and a strong dynamic transformation from perception to sustained internal simulation (PC3+) **(Fig 5A-B)**. The AVP-induced stabilization in this state may reflect a proactive engagement with the internal threat representation and may reflect that AVP enhances an internal ‘threat simulation’ mode underlying an active threat evaluation and leading to a subjectively elongated perception of time [79] and enhanced arousal [80]. The shift in the processing mode (partly) aligns with models describing adaptive decision-making under uncertainty [71].

#### Losartan Induces a more Controlled Processing Mode

LT reduced subjective state anxiety in the absence of changes in peripheral cardiovascular activity. This pattern may reflect a – previously described - delayed peak efficacy of losartan on cardiovascular function in normotensive subjects [20,21] and the differentiation of systems underlying subjective experience and autonomic reactivity or threats [81,82]. While the central blood-brain barrier penetration of losartan remains debated our findings are in line with previously reported effects in human psychopharmacological studies and models suggesting direct effects via blood-brain barrier crossing and/or via actions on circumventricular organs and subsequent modulation of central autonomic and stress circuits, or peripheral effects mediating the anxiolytic responses [20,21,83–85].

Behaviorally, LT led to temporal compression (*β* < PLC), indicating accelerated decision-making. Pupillometrically, LT reduced overall arousal (smaller pupil responses) while preserving post-stimulus threat discrimination— but only at high velocities, possible reflecting a selective, efficient allocation of resources to imminent threats.

HMM further qualified these effects such that LT: (1) resulted in the highest sequential entropy, indicating a highly flexible and unpredictable processing stream devoid of rigid state sequences, and, (2) specifically suppressed transitions into the high-anxiety, high-simulation L2 state (+,+,+) that is characteristic of the AVP-driven response. Notably, while both LT and AVP increased the steady-state probability of the proactive preparation state L3 (-,+,+), the direct opposite of the PLC-dominant E1(+,-,-), LT’s critical action might have been to prevent maintaining in a hyper-vigilant state of L2.

Collectively, these results demonstrate that LT induces a more controlled mode, rather than promoting a specific alternative profile. As such LT’s threat modulation may be based on preventing fixed responses [86] and enhancing flexibile proactive preparation (as reflected in the increased steady-state probability of L3). Together this may promote efficient resource allocation, accelerated time-perception (temporal compression), and an overall reduction in subjective anxiety.

#### Commonalities of neuropeptide threat modulation

Despite the differences both neuropeptides showed a common effect of heightened pupil response to threat versus non-threat stimuli specifically during the post-stimulus imaginary phase, a modulation that was absent under placebo. This indicates that they modulate the internal evaluation of threat in addition to processes driven by sensory input. AVP may operate through potentiation and stabilizing a hyper-vigilant state (L2) leading to exaggerated threat engagement and subjective time dilation. In contrast, blockade of the renin-angiotensin II system may promote cognitive flexibility (highest entropy) by specifically suppressing transitions into the L2 state promoting efficient threat processing and subjective time compression. These distinct modulatory roles may render the systems as promising and distinct therapeutic targets, such that LT may alleviate anxiety, affective and hyperarousal (see also [85,87]) while AVP may enhance threat sensitivity.

### Limitations

Our study has several limitations. First, while we analyzed pupillary and behavioral responses, we did not examine their link to neural activation patterns. Future work could combine pupillometry with neuroimaging to directly map these relationships. Second, although chromatic differences existed between stimulus phases (colored vs. grayscale), our key findings—such as threat-specific pupillary dilation and pharmacological modulation—were consistent across matched conditions, suggesting cognitive-emotional rather than light reflex. Nevertheless, strictly controlled visual properties in future designs would strengthen validity. Finally, our principal component interpretations, while aligned with theoretical frameworks (e.g., arousal modulation, threat detection networks), remain inferential. Empirical validation of these components’ neural substrates is needed.

## Supporting information

Complete Supplementary Materials

## Data Availability

All data produced in the present study are available upon reasonable request to the authors

## Abbreviations

LT: losartan;
PLC: placebo;
AVP: arginine vasopressin;
RAS: renin-angiotensin system;
LC-NE: locus coeruleus-norepinephrine system;
aTTC: actual time to collision;
jTTC: judged time to collision;
PSV: physical stimulus velocity;
SBP: systolic blood pressure;
DBP: diastolic blood pressure;
Early/Late Imag: early/late imagination phase;
FLMM: functional linear mixed model;
FPCA: functional principal component analysis;
PC: principal component;
HMM: hidden Markov model

## Data Availability

All group-level data (including judged Time-To-Collision, anxiety ratings, and blood pressure measurements) and normalized pupil data will be made publicly available upon acceptance in the Open Science Framework (OSF) repository (DOI will be provided upon publication).

## References

1. Gao N, Wang H, Xu X, Yang Z, Zhang T. Angiotensin II induces cognitive decline and anxiety-like behavior via disturbing pattern of theta-gamma oscillations. Brain Res Bull. 2021;174: 84–91. doi:10.1016/j.brainresbull.2021.06.002

2. Liu X, Jiao G, Zhou F, Kendrick KM, Yao D, Gong Q, et al. A neural signature for the subjective experience of threat anticipation under uncertainty. Nat Commun. 2024;15: 1544. doi:10.1038/s41467-024-45433-6

3. Taschereau-Dumouchel V, Kawato M, Lau H. Multivoxel pattern analysis reveals dissociations between subjective fear and its physiological correlates. Mol Psychiatry. 2020;25: 2342–2354. doi:10.1038/s41380-019-0520-3

4. Thieu MK, Ayzenberg V, Lourenco SF, Kragel PA. Visual looming is a primitive for human emotion. iScience. 2024;27: 109886. doi:10.1016/j.isci.2024.109886

5. Zhou F, Geng Y, Xin F, Li J, Feng P, Liu C, et al. Human extinction learning is accelerated by an angiotensin antagonist via ventromedial prefrontal cortex and its connections with basolateral amygdala. Biol Psychiatry. 2019;86: 910–920. doi:10.1016/j.biopsych.2019.07.007

6. Ohman A. The role of the amygdala in human fear: Automatic detection of threat. Psychoneuroendocrinology. 2005;30: 953–958. doi:10.1016/j.psyneuen.2005.03.019

7. Adolphs R. Neural systems for recognizing emotion. Curr Opin Neurobiol. 2002;12: 169–177. doi:10.1016/s0959-4388(02)00301-x

8. LeDoux JE, Brown R. A higher-order theory of emotional consciousness. Proceedings of the National Academy of Sciences. 2017;114: E2016–E2025. doi:10.1073/pnas.1619316114

9. Card G, Dickinson M. Performance trade-offs in the flight initiation of drosophila. J Exp Biol. 2008;211: 341–353. doi:10.1242/jeb.012682

10. Schiff W. Perception of impending collision: A study of visually directed avoidant behavior. Psychological Monographs: General and Applied. 1965;79: 1–26. doi:10.1037/h0093887

11. Yilmaz M, Meister M. Rapid innate defensive responses of mice to looming visual stimuli. Current Biology. 2013;23: 2011–2015. doi:10.1016/j.cub.2013.08.015

12. Cooper WE, Samia DSM, Blumstein DT. Chapter five - FEAR, spontaneity, and artifact in economic escape theory: A review and prospectus. In: Naguib M, Brockmann HJ, Mitani JC, Simmons LW, Barrett L, Healy S, et al., editors. Advances in the Study of Behavior. Academic Press; 2015. pp. 147–179. doi:10.1016/bs.asb.2015.02.002

13. Vagnoni E, Lourenco SF, Longo MR. Threat modulates perception of looming visual stimuli. Current Biology. 2012;22: R826–R827. doi:10.1016/j.cub.2012.07.053

14. Cole S, Balcetis E, Dunning D. Affective signals of threat increase perceived proximity. Psychol Sci. 2013;24: 34–40. doi:10.1177/0956797612446953

15. Brendel E, Hecht H, DeLucia PR, Gamer M. Emotional effects on time-to-contact judgments: Arousal, threat, and fear of spiders modulate the effect of pictorial content. Exp Brain Res. 2014;232: 2337–2347. doi:10.1007/s00221-014-3930-0

16. Huber D, Veinante P, Stoop R. Vasopressin and oxytocin excite distinct neuronal populations in the central amygdala. Science. 2005;308: 245–248. doi:10.1126/science.1105636

17. Meyer-Lindenberg A, Domes G, Kirsch P, Heinrichs M. Oxytocin and vasopressin in the human brain: Social neuropeptides for translational medicine. Nat Rev Neurosci. 2011;12: 524–538. doi:10.1038/nrn3044

18. Thompson RR, George K, Walton JC, Orr SP, Benson J. Sex-specific influences of vasopressin on human social communication. Proc Natl Acad Sci U S A. 2006;103: 7889–7894. doi:10.1073/pnas.0600406103

19. Reinecke A, Browning M, Klein Breteler J, Kappelmann N, Ressler KJ, Harmer CJ, et al. Angiotensin regulation of amygdala response to threat in high-trait-anxiety individuals. Biol Psychiatry Cogn Neurosci Neuroimaging. 2018;3: 826–835. doi:10.1016/j.bpsc.2018.05.007

20. Zhang R, Zhao W, Qi Z, Xu T, Zhou F, Becker B. Angiotensin II regulates the neural expression of subjective fear in humans: A precision pharmaco-neuroimaging approach. Biol Psychiatry Cogn Neurosci Neuroimaging. 2023;8: 262–270. doi:10.1016/j.bpsc.2022.09.008

21. Xu T, Zhou X, Jiao G, Zeng Y, Zhao W, Li J, et al. Angiotensin antagonist inhibits preferential negative memory encoding via decreasing hippocampus activation and its coupling with the amygdala. Biological Psychiatry: Cognitive Neuroscience and Neuroimaging. 2022;7: 970–978. doi:10.1016/j.bpsc.2022.05.007

22. Szczepanska-Sadowska E. Role of neuropeptides in central control of cardiovascular responses to stress. J Physiol Pharmacol. 2008;59 Suppl 8: 61–89.

23. Höhle S, Culman J, Boser M, Qadri F, Unger T. Effect of angiotensin AT2 and muscarinic receptor blockade on osmotically induced vasopressin release. Eur J Pharmacol. 1996;300: 119–123. doi:10.1016/0014-2999(95)00855-1

24. von Bohlen und Halbach O, Albrecht D. The CNS renin-angiotensin system. Cell Tissue Res. 2006;326: 599–616. doi:10.1007/s00441-006-0190-8

25. Guo F, Zou J, Wang Y, Fang B, Zhou H, Wang D, et al. Human subcortical pathways automatically detect collision trajectory without attention and awareness. PLOS Biology. 2024;22: e3002375. doi:10.1371/journal.pbio.3002375

26. Armstrong T, Bilsky SA, Zhao M, Olatunji BO. Dwelling on potential threat cues: An eye movement marker for combat-related ptsd. Depression and Anxiety. 2013;30: 497–502. doi:10.1002/da.22115

27. Chen L, Yuan X, Xu Q, Wang Y, Jiang Y. Subliminal impending collision increases perceived object size and enhances pupillary light reflex. Front Psychol. 2016;7. doi:10.3389/fpsyg.2016.01897

28. Mathôt S, Van der Stigchel S. New light on the mind’s eye: The pupillary light response as active vision. Curr Dir Psychol Sci. 2015;24: 374–378. doi:10.1177/0963721415593725

29. Joshi S, Li Y, Kalwani RM, Gold JI. Relationships between pupil diameter and neuronal activity in the locus coeruleus, colliculi, and cingulate cortex. Neuron. 2016;89: 221–234. doi:10.1016/j.neuron.2015.11.028

30. Hernández-Pérez OR, Hernández VS, Nava-Kopp AT, Barrio RA, Seifi M, Swinny JD, et al. A Synaptically Connected Hypothalamic Magnocellular Vasopressin-Locus Coeruleus Neuronal Circuit and Its Plasticity in Response to Emotional and Physiological Stress. Front Neurosci. 2019;13. doi:10.3389/fnins.2019.00196

31. Gong W, Lü J, Wang F, Wang B, Wang M, Huang H. Effects of angiotensin type 2 receptor on secretion of the locus coeruleus in stress-induced hypertension rats. Brain Research Bulletin. 2015;111: 62–68. doi:10.1016/j.brainresbull.2014.12.011

32. Mobbs D, Hagan CC, Dalgleish T, Silston B, Prévost C. The ecology of human fear: Survival optimization and the nervous system. Front Neurosci. 2015;9. doi:10.3389/fnins.2015.00055

33. Mineka S, Öhman A. Phobias and preparedness: The selective, automatic, and encapsulated nature of fear. Biological Psychiatry. 2002;52: 927–937. doi:10.1016/S0006-3223(02)01669-4

34. Phelps EA. Emotion and cognition: Insights from studies of the human amygdala. Annual Review of Psychology. 2006;57: 27–53. doi:10.1146/annurev.psych.56.091103.070234

35. Watson D, Clark LA, Tellegen A. Development and validation of brief measures of positive and negative affect: The PANAS scales. Journal of Personality and Social Psychology. 1988;54: 1063–1070. doi:10.1037/0022-3514.54.6.1063

36. Spielberger CD. State-trait anxiety inventory for adults. 2012. doi:10.1037/t06496-000

37. Heimberg RG, Horner KJ, Juster HR, Safren SA, Brown EJ, Schneier FR, et al. Psychometric properties of the liebowitz social anxiety scale. Psychological Medicine. 1999;29: 199–212. doi:10.1017/S0033291798007879

38. Vetter RS. Calling all arachnophobic entomologists: A request for information. American Entomologist. 2012;58: 199–201. doi:10.1093/ae/58.4.199

39. Zhuang Q, Zheng X, Becker B, Lei W, Xu X, Kendrick KM. Intranasal vasopressin like oxytocin increases social attention by influencing top-down control, but additionally enhances bottom-up control. Psychoneuroendocrinology. 2021;133: 105412. doi:10.1016/j.psyneuen.2021.105412

40. Kou J, Zhang Y, Zhou F, Sindermann C, Montag C, Becker B, et al. A randomized trial shows dose-frequency and genotype may determine the therapeutic efficacy of intranasal oxytocin. Psychological Medicine. 2022;52: 1959–1968. doi:10.1017/S0033291720003803

41. Quintana DS, Smerud KT, Andreassen OA, Djupesland PG. Evidence for intranasal oxytocin delivery to the brain: Recent advances and future perspectives. Therapeutic Delivery. 2018 [cited 5 Apr 2025]. doi:10.4155/tde-2018-0002

42. Berger A, Kiefer M. Comparison of different response time outlier exclusion methods: A simulation study. Front Psychol. 2021;12. doi:10.3389/fpsyg.2021.675558

43. Ratcliff R. Methods for dealing with reaction time outliers. Psychological Bulletin. 1993;114: 510–532. doi:10.1037/0033-2909.114.3.510

44. Hayes TR, Petrov AA. Mapping and correcting the influence of gaze position on pupil size measurements. Behav Res. 2016;48: 510–527. doi:10.3758/s13428-015-0588-x

45. Kret ME, Sjak-Shie EE. Preprocessing pupil size data: Guidelines and code. Behav Res. 2019;51: 1336–1342. doi:10.3758/s13428-018-1075-y

46. Eagleman DM. Human time perception and its illusions. Curr Opin Neurobiol. 2008;18: 131–136. doi:10.1016/j.conb.2008.06.002

47. Burnham KP, Anderson DR. Multimodel inference. Sociological Methods & Research. 2004 [cited 30 Sept 2025]. doi:10.1177/0049124104268644

48. Laeng B, Sulutvedt U. The eye pupil adjusts to imaginary light. Psychological Science. 2013 [cited 3 Apr 2025]. doi:10.1177/0956797613503556

49. Eilers PHC, Marx BD. Flexible smoothing with B-splines and penalties. Statistical Science. 1996;11: 89–121. doi:10.1214/ss/1038425655

50. Dan EL, Dînşoreanu M, Mureşan RC. Accuracy of six interpolation methods applied on pupil diameter data. 2020 IEEE International Conference on Automation, Quality and Testing, Robotics (AQTR). 2020. pp. 1–5. doi:10.1109/AQTR49680.2020.9129915

51. Hershman R, Henik A, Cohen N. A novel blink detection method based on pupillometry noise. Behav Res. 2018;50: 107–114. doi:10.3758/s13428-017-1008-1

52. Greven S, Scheipl F. A general framework for functional regression modelling. Statistical Modelling. 2017 [cited 30 Sept 2025]. doi:10.1177/1471082X16681317

53. Bates D, Kliegl R, Vasishth S, Baayen H. Parsimonious mixed models. arXiv; 2018. doi:10.48550/arXiv.1506.04967

54. Viviani R, Grön G, Spitzer M. Functional principal component analysis of fMRI data. Hum Brain Mapp. 2005;24: 109–129. doi:10.1002/hbm.20074

55. Caliński T, Harabasz J. A dendrite method for cluster analysis. Communications in Statistics - Theory and Methods. 1974 [cited 30 Sept 2025]. Available: https://www.tandfonline.com/doi/abs/10.1080/03610927408827101

56. Vidaurre D, Smith SM, Woolrich MW. Brain network dynamics are hierarchically organized in time. Proceedings of the National Academy of Sciences. 2017;114: 12827–12832. doi:10.1073/pnas.1705120114

57. Rabiner LR. A tutorial on hidden markov models and selected applications in speech recognition. Proceedings of the IEEE. 1989;77: 257–286. doi:10.1109/5.18626

58. Renner LF, Włodarczak M. When a dog is a cat and how it changes your pupil size: Pupil dilation in response to information mismatch. Interspeech 2017. ISCA; 2017. pp. 674–678. doi:10.21437/Interspeech.2017-353

59. Benjamini Y, Hochberg Y. Controlling the false discovery rate: A practical and powerful approach to multiple testing. Journal of the Royal Statistical Society Series B (Methodological). 1995;57: 289–300.

60. Jazayeri M, Shadlen MN. Temporal context calibrates interval timing. Nat Neurosci. 2010;13: 1020–1026. doi:10.1038/nn.2590

61. Ratcliff R, McKoon G. The diffusion decision model: Theory and data for two-choice decision tasks. Neural Comput. 2008;20: 873–922. doi:10.1162/neco.2008.12-06-420

62. Bogacz R, Brown E, Moehlis J, Holmes P, Cohen JD. The physics of optimal decision making: A formal analysis of models of performance in two-alternative forced-choice tasks. Psychol Rev. 2006;113: 700–765. doi:10.1037/0033-295X.113.4.700

63. Gold JI, Shadlen MN. The neural basis of decision making. Annu Rev Neurosci. 2007;30: 535–574. doi:10.1146/annurev.neuro.29.051605.113038

64. Mathôt S, Fabius J, Van Heusden E, Van der Stigchel S. Safe and sensible preprocessing and baseline correction of pupil-size data. Behav Res. 2018;50: 94–106. doi:10.3758/s13428-017-1007-2

65. (PDF) THE PUPIL: ANATOMY, PHYSIOLOGY, AND CLINICAL APPLICATIONS: by irene E. Loewenfeld. 1999. Oxford: butterworth-heinemann. Price pound180. Pp. 2278. ISBN 0-750-67143-2. ResearchGate. 2024 [cited 11 Apr 2025]. doi:10.1093/brain/124.9.1881

66. Henderson RR, Bradley MM, Lang PJ. Emotional imagery and pupil diameter. Psychophysiology. 2018;55: e13050. doi:10.1111/psyp.13050

67. Laeng B, Sulutvedt U. The eye pupil adjusts to imaginary light. Psychol Sci. 2014;25: 188–197. doi:10.1177/0956797613503556

68. Mobbs D, Headley DB, Ding W, Dayan P. Space, time, and fear: Survival computations along defensive circuits. Trends Cogn Sci. 2020;24: 228–241. doi:10.1016/j.tics.2019.12.016

69. Bradley MM, Miccoli L, Escrig MA, Lang PJ. The pupil as a measure of emotional arousal and autonomic activation. Psychophysiology. 2008;45: 602–607. doi:10.1111/j.1469-8986.2008.00654.x

70. Wang C-A, Munoz DP. Modulation of stimulus contrast on the human pupil orienting response. Eur J Neurosci. 2014;40: 2822–2832. doi:10.1111/ejn.12641

71. Aston-Jones G, Cohen JD. An integrative theory of locus coeruleus-norepinephrine function: Adaptive gain and optimal performance. Annu Rev Neurosci. 2005;28: 403–450. doi:10.1146/annurev.neuro.28.061604.135709

72. Mobbs D, Marchant JL, Hassabis D, Seymour B, Tan G, Gray M, et al. From threat to fear: The neural organization of defensive fear systems in humans. J Neurosci. 2009;29: 12236–12243. doi:10.1523/JNEUROSCI.2378-09.2009

73. Cross CP, Campbell A. Women’s aggression. Aggression and Violent Behavior. 2011;16: 390–398. doi:10.1016/j.avb.2011.02.012

74. Knyazev GG, Bocharov AV, Slobodskoj-Plusnin JY. Hostility- and gender-related differences in oscillatory responses to emotional facial expressions. Aggress Behav. 2009;35: 502–513. doi:10.1002/ab.20318

75. Partala T, Surakka V. Pupil size variation as an indication of affective processing. International Journal of Human-Computer Studies. 2003;59: 185–198. doi:10.1016/S1071-5819(03)00017-X

76. Kou J, Lan C, Zhang Y, Wang Q, Zhou F, Zhao Z, et al. In the nose or on the tongue? Contrasting motivational effects of oral and intranasal oxytocin on arousal and reward during social processing. Transl Psychiatry. 2021;11: 94. doi:10.1038/s41398-021-01241-w

77. Le J, Zhang L, Zhao W, Zhu S, Lan C, Kou J, et al. Infrequent intranasal oxytocin followed by positive social interaction improves symptoms in autistic children: A pilot randomized clinical trial. Psychother Psychosom. 2022;91: 335–347. doi:10.1159/000524543

78. Zhuang Q, Zheng X, Yao S, Zhao W, Becker B, Xu X, et al. Oral administration of oxytocin, like intranasal administration, decreases top-down social attention. Int J Neuropsychopharmacol. 2022;25: 912–923. doi:10.1093/ijnp/pyac059

79. Droit-Volet S. Time perception, emotions and mood disorders. Journal of Physiology-Paris. 2013;107: 255–264. doi:10.1016/j.jphysparis.2013.03.005

80. Neumann ID, Landgraf R. Balance of brain oxytocin and vasopressin: Implications for anxiety, depression, and social behaviors. Trends in Neurosciences. 2012;35: 649–659. doi:10.1016/j.tins.2012.08.004

81. Liu X, Jiao G, Zhou F, Kendrick KM, Yao D, Gong Q, et al. A neural signature for the subjective experience of threat anticipation under uncertainty. Nat Commun. 2024;15: 1544. doi:10.1038/s41467-024-45433-6

82. Zhang R, Gan X, Xu T, Yu F, Wang L, Song X, et al. A neurofunctional signature of affective arousal generalizes across valence domains and distinguishes subjective experience from autonomic reactivity. Nat Commun. 2025;16: 6492. doi:10.1038/s41467-025-61706-0

83. Salim H, Jones AM. Angiotensin II receptor blockers (ARBs) and manufacturing contamination: A retrospective national register study into suspected associated adverse drug reactions. Br J Clin Pharmacol. 2022;88: 4812–4827. doi:10.1111/bcp.15411

84. Pulcu E, Shkreli L, Holst CG, Woud ML, Craske MG, Browning M, et al. The effects of the angiotensin II receptor antagonist losartan on appetitive versus aversive learning: A randomized controlled trial. Biol Psychiatry. 2019;86: 397–404. doi:10.1016/j.biopsych.2019.04.010

85. The brain renin angiotensin system: A novel precision target for neurofunctional symptom regulation | request PDF. In: ResearchGate [Internet]. 25 Aug 2025 [cited 29 Sept 2025]. doi:10.31234/osf.io/fbcv4_v1

86. Gillan CM, Apergis-Schoute AM, Morein-Zamir S, Urcelay GP, Sule A, Fineberg NA, et al. Functional neuroimaging of avoidance habits in obsessive-compulsive disorder. AJP. 2015;172: 284–293. doi:10.1176/appi.ajp.2014.14040525

87. Angiotensin II regulates the neural expression of subjective fear in humans: A precision pharmaco-neuroimaging approach. Biological Psychiatry: Cognitive Neuroscience and Neuroimaging. 2023;8: 262–270. doi:10.1016/j.bpsc.2022.09.008

