## Supplementary material for "Vasopressin and angiotensin II differentially modulate human fear response dynamics to looming threats": Complete Supplementary Materials

**Consort**

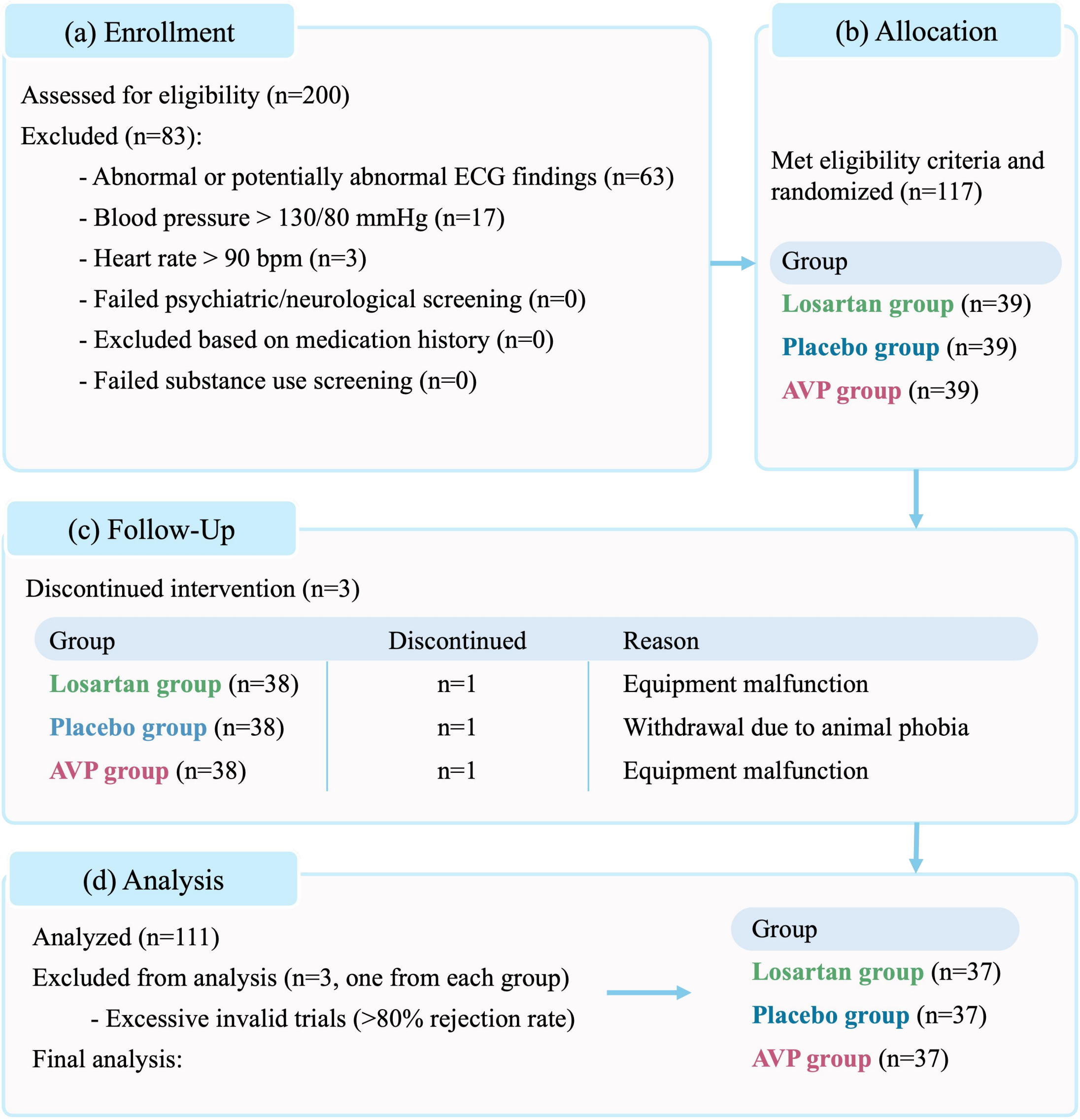


**S1 Fig. Participant screening and randomization flow chart.**

### Supplementary Method

#### S1 Detailed Participant Screening Procedures

Prior to participation, all volunteers underwent an electrocardiogram (ECG) examination to rule out potential cardiovascular contraindications. To minimize potential confounds from psychoactive substances, participants were required to abstain from caffeine and alcohol for 24 hours prior to the testing session.

For female participants, a urine-based pregnancy test was administered on the day of the experiment to exclude pregnancy. Additionally, the phase of their menstrual cycle was documented based on self-report following established procedures [37,38] to account for potential hormonal influences.

#### S2 Drug Preparation and Administration Protocol

Argipressin (AVP; Bio-Techne China Co., Ltd) was aliquoted and stored at -20° C until the day of the experiment. For administration, AVP was dissolved in a sterile vehicle solution of saline and glycerol, passed through a 0.22 μm Millipore filter to ensure sterility, and loaded into spray bottles. Placebo sprays were prepared in an identical manner but omitted the active AVP peptide.

Following our validated protocol (Kou et al., 2021; Zhuang et al., 2022), each participant received six separate 0.1 ml puffs (total volume = 0.6 ml), with a 30-second retention period between puffs. The puffs were administered in an alternating fashion: three puffs were directed to the superior surface of the tongue and three to the inferior surface. Participants were instructed to avoid swallowing for the duration of the retention period.

#### S3 Stimulus Preparation and Presentation

Stimulus images were normalized to a size of 400 × 250 pixels against a uniform grey background using Adobe Photoshop. The experiment was programmed and presented using Psychtoolbox-3 running under MATLAB (R2018b).

#### S4 Data Quality Control and Temporal Normalization

To ensure the validity of our pupillometry analysis, trials were subjected to predefined quality control criteria prior to inclusion. These criteria were designed to remove trials with fundamental data integrity issues while aligning with established practices in pupillometry research (Mathôt et al., 2018). The application of these criteria resulted in the exclusion of 9.65% of trials (1,506 trials), yielding 14,266 valid trials for analysis. This exclusion rate is well within the expected and acceptable range of 10 – 20% for pupillometry studies (Hayes & Petrov, 2016; Winn et al., 1994). Critically, the number of excluded trials did not differ between treatment groups ($\chi^2$ (2) = 0.46, $p$ = .53), arguing against any biasing effects of the exclusion criteria on our results.

Trials were excluded based on the following principled criteria:

1. **Missing Behavioral Responses:** Trials without a recorded behavioral response were excluded, as they preclude the analysis of time-to-collision judgments.

2. **Insufficient Valid Data Points:** Trials with fewer than 10 overall valid data points, or fewer than 3 valid points within any predefined analysis phase (baseline, stimulus presentation, or imagination), were excluded. Given our 100 Hz sampling rate, the phase-specific requirement of 3 points translates to a lenient threshold of only 30 ms of valid data. This ensures we selectively excluded only trials with extreme artifacts (e.g., complete signal loss from prolonged blinks) while retaining the majority of data (Hershman et al., 2018; Kret & De Dreu, 2019).

3. **Short Trial Duration (< 2300 ms)**: This criterion was informed by the minimum data requirement for our event-locked and time-normalized analyses. The calculation is as follows: 300 ms (baseline) + 1000 ms (stimulus presentation) + 1000 ms (minimum imagination period) = 2300 ms. This 1000 ms post-stimulus window was critical to cleanly extract the late imagination epoch (the final 500 ms before response) without truncation from an early response. Including shorter trials would introduce inconsistency and potential bias.

4. **Poor Signal Integrity:** Signal integrity was verified through temporal continuity checks and autocorrelation analysis to identify and eliminate trials with pervasive, uncorrectable high-frequency noise, ensuring results are not driven by artifact-laden data (Mathôt et al., 2018).

For subsequent analysis of the variable-duration imagination periods, we implemented a piecewise linear time warping procedure to normalize temporal dynamics across trials, a common approach for handling temporal misalignment in functional data (Ramsay & Silverman, 1997). The approach preserved the functional segregation between stimulus processing and imagination phases by mapping the fixed stimulus presentation epoch (300-1300 ms) to the normalized interval [0, 0.5], while the variable imagination epoch (from 1300 ms to response) was mapped to [0.5, 1.0]. Each normalized epoch was resampled at 100 equidistant points, creating a standardized 200-point representation per trial that maintained the relative temporal structure within phases while enabling direct comparison across trials. Finally, the normalized time series were smoothed using cubic B-spline functions (order 3, 4 basis components) to reduce high-frequency noise while preserving authentic temporal dynamics in the pupillary response (Dan et al., 2020; Eilers & Marx, 1996).

### Supplementary Results

**S1 Table. Demographics and questionnaire scores between treatment groups.**

| Variables | LT | PLC | AVP | *p*-value |
|---|---|---|---|---|
| Age, Years | 21.92±2.28 | 22.26±2.52 | 21.63±2.07 | .49 |
| PANAS-Positive | 29.50±4.70 | 27.47±5.44 | 27.55±6.44 | .21 |
| PANAS-Negative | 15.39±5.14 | 16.39±5.14 | 14.79±5.87 | .43 |
| LSAS-Anxiety | 47.29±12.17 | 45.47±8.62 | 43.47±9.29 | .27 |
| LSAS-Avoidance | 43.84±11.93 | 43.87±10.21 | 41.34±8.60 | .48 |
| TAI | 37.66±7.63 | 39.69±6.29 | 37.16±8.32 | .31 |
| SAI-Baseline | 35.29±8.19 | 37.58±6.67 | 35.39±7.67 | .34 |
| SAI-Post | 34.21±8.29 | 37.81±8.16 | 34.32±9.45 | .13 |
| AFQ-Rabbit | 6.29±1.31 | 6.58±1.42 | 6.37±1.89 | .71 |
| AFQ-Butterfly | 7.82±3.54 | 8.36±3.35 | 7.82±2.91 | .71 |
| AFQ-Snake | 19.00±4.67 | 17.72±5.49 | 18.08±4.96 | .53 |
| AFQ-Spider | 19.71±5.04 | 19.81±4.64 | 19.05±5.33 | .78 |

Values are presented as mean ± SD. The *df* for *F* values is 108.

PANAS, Positive and Negative Affect Schedule; LSAS, Liebowitz Social Anxiety Scale; TAI and SAI, Trait and State subscales of the State-Trait Anxiety Inventory; AFQ, Animal Fear Questionnaire; LT, losartan; PLC, placebo; AVP: vasopressin.

### Supplementary Results: Functional Principal Components

Functional Principal Component Analysis (FPCA) of pupillary dynamics revealed three dominant components (eigenfunctions) accounting for 87.51% of total variance. As shown in Fig. 4A, their temporal weight functions displayed distinct patterns across the stimulus approach (0-0.5 s) and post-stimulus imagination (0.5-1.0 s) phases: PC1 (62.7%) showed positive weights that rose during stimulus approach and plateaued during imagination; PC2 (17.6%) transitioned from negative weights during approach to positive weights during imagination; and PC3 (7.3%)

exhibited a positive-negative-positive oscillation spanning both phases.

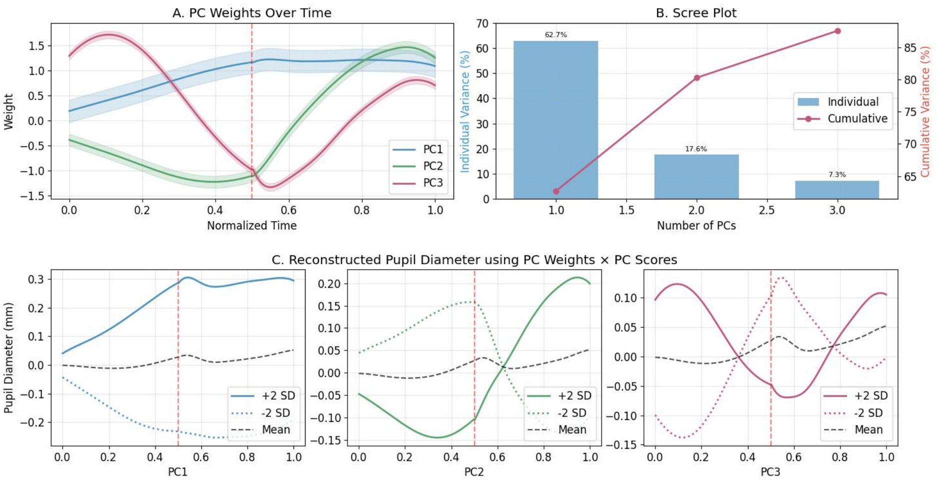


**S2 Fig. Principal component analysis of pupillary features. (A)** Temporal weight functions showing the contribution of PC1-3 over normalized time. The solid lines represent the mean weights, with shaded areas indicating the 95% confidence intervals. The vertical dashed red line marks the stimulus onset. **(B)** Scree plot displaying both individual (blue bars) and cumulative (red line) explained variance for the first three PCs. PC1, PC2, and PC3 account for 62.7%, 17.6%, and 7.3% of the total variance respectively, with a cumulative explained variance of 87.6%. **(C)** Reconstructed pupil diameter changes based on PC weights × PC scores for PC1-3. For each PC, the solid line represents pupil diameter changes at +2 SD of PC scores, the dotted line at -2 SD, and the dashed line shows the mean response. Changes in pupil diameter are shown in millimeters (mm).

#### Effects of Experimental Conditions on Principal Components

Mixed ANOVAs on the principal components (PC1-3) revealed distinct effects of experimental conditions (Fig. 5).

All components showed significant PSV main effects (PC1: $F = 105$, $p < .001$, $\eta_p^2 = .40$; PC2: $F = 105$, $p < .001$, $\eta_p^2 = .31$; PC3: $F = 7.32$, $p = .005$, $\eta_p^2 = 0.63$), each with distinct patterns: extreme speeds (V1, V5) elicited higher PC1 scores but lower PC3 scores than medium speeds (V2-V4), while PC2 showed progressively increased scores with speed (Fig. 5C).

For PC1 (sustained dilation pattern), we found a significant Treatment × Sex × Isthreaten interaction ($F = 3.37$, $p = .023$, $\eta_p^2 = 0.69$). Females showed treatment-dependent responses to threat: under non-threatening conditions, the PLC group exhibited higher PC1 scores than drug groups (*p*s < .01), while under threatening conditions, both PLC and AVP groups showed higher scores than the LT group (*p*s < .05); Males showed no treatment effects across conditions (*p*s > .48) (Fig. 5A1, A2).

PC2 (biphasic pattern) demonstrated significant main effects of Treatment ($F = 7.82$, $p < .001$, $\eta_p^2 = 0.30$) and Isthreaten ($F = 9.76$, $p = .002$, $\eta_p^2 = 0.24$). The AVP group showed higher PC2 scores compared to other groups (AVP vs PLC: $p = .006$; AVP vs LT: $p = .025$), whereas LT-PLC showed no difference ($p = .53$). Threatening stimuli elicited higher scores than non-threatening ones ($p < .001$). A significant Treatment × Isthreaten interaction revealed greater responses to threatening versus non-threatening stimuli in both drug groups (*p*s < .01), but not in PLC ($p = .45$) (Fig. 5B).

PC3 (triphasic pattern) showed a significant main effect of Isthreaten ($F = 8.12$, $p = .005$, $\eta_p^2 = 0.22$), with threatening stimuli eliciting higher scores than non-threatening ones.

**Relationships Between Principal Components and Individual Difference Variables**

We examined correlations between pupillary response components and individual differences in anxiety changes (Post-Pre State Anxiety scores) and judged time-to-collision (jTTC).

The most notable finding was a moderate negative correlation between PC2 (biphasic pattern) and anxiety change scores ($r = -0.302$, $p = .001$) (Fig 5D). Further analysis comparing high-anxiety (top 20%) versus low-anxiety (bottom 20%) groups revealed significantly lower PC2 scores in the high-anxiety group (M = -0.025, SD = 0.035) compared to the low-anxiety group (M = 0.010, SD = 0.03) ($t = 3.22$, $p = .003$, Cohen's $d = 1.09$). This suggests that individuals with increased anxiety showed stronger pupil dilation during stimulus presentation but enhanced constriction after stimulus offset and provides a behavioral link of the PC2 to subjective anxiety.

For behavioral responses, PC3 (triphasic pattern) showed a significant positive correlation with jTTC ($r = 0.263$, $p = .004$), indicating that slower responders exhibited a distinctive pattern of weakened dilation followed by enhanced constriction-dilation cycles. PC1 showed a marginal negative trend with jTTC ($r = -0.169$, $p = .069$), while PC2's correlation with jTTC was non-significant ($r = 0.147$, $p = .114$) (Fig 5E).

**Associations Between FPCA-derived Components and Phase-specific Pupillary Features**

PC1 showed strong positive correlations with pupil dilation state ($r = 0.615$) and change rate ($r = 0.650$) during stimulus presentation, while negatively correlating with baseline pupil diameter ($r = -0.177$), suggesting greater dilation potential in participants with smaller baseline pupils.

PC2 demonstrated strong positive correlations with post-stimulus pupil dilation state ($r = 0.668$) and change rate ($r = 0.603$), while showing a moderate negative correlation with during-stimulus pupil state ($r = -0.352$), capturing the inhibition-to-activation transition.

PC3 exhibited strong negative correlation with during-stimulus change rate ($r = -0.580$) and a moderate positive correlation with post-stimulus change rate ($r = 0.337$), reflecting temporal dynamic regulation (Fig 5F). See Supplementary Materials for feature calculation details.

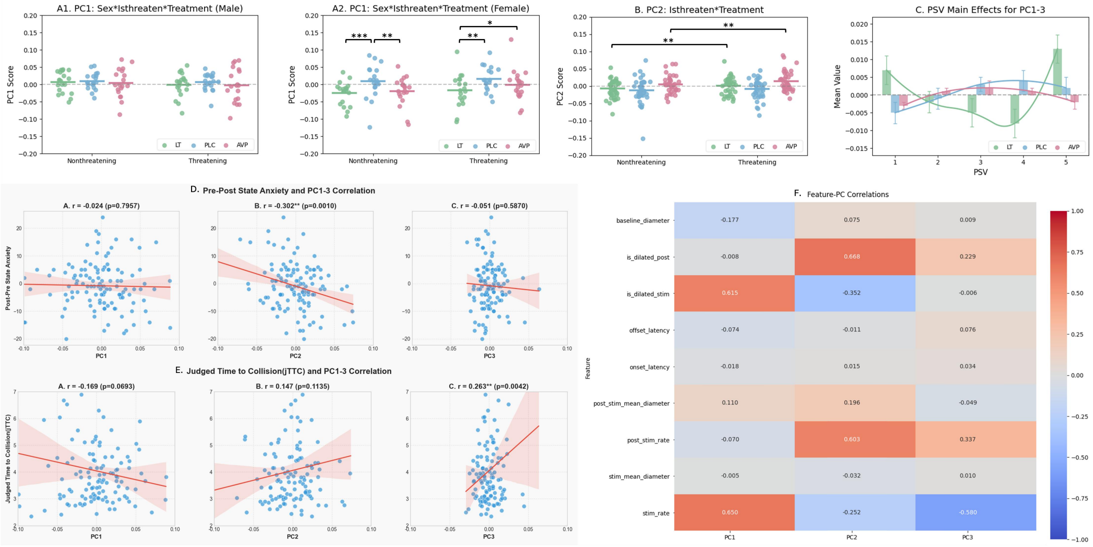


**S3 Fig. Relationships between pupillary principal components, experimental conditions, and individual differences.** **(A)** Effects of sex, treatment, and threat conditions on PC1 scores: **(A1)** No significant differences among treatments in males; **(A2)** In females, PLC group showed higher PC1 scores than drug groups under non-threatening conditions, while PLC and AVP groups showed higher scores than LT group under threatening conditions. **(B)** PC2 scores across conditions: AVP group significantly higher than PLC and LT groups, and threatening stimuli higher than non-threatening stimuli; AVP and LT groups showed increased PC2 scores under threat, while PLC group remained unchanged. **(C)** Principal component scores across different speeds: PC1 sensitive to extreme speeds (V1,V5), PC2 increased with speed, PC3 more sensitive to medium speeds (V2-V4). **(D)** Negative correlation between PC2 and state anxiety change (Post-Pre) ($r$=-0.302, $p$=.001). **(E)** Positive correlation between PC3 and judged time-to-collision (jTTC) ($r$=0.263, $p$<.01). **(F)** Correlations between principal components and pupillary parameters: PC1 positively correlated with pupil dilation during stimulus presentation, PC2 positively correlated with post-stimulus pupillary changes but negatively with during-stimulus pupil state, PC3 negatively correlated with during-stimulus change rate ($r$=-0.580) and positively with post-stimulus change rate ($r$=0.337). **$p$<.01.

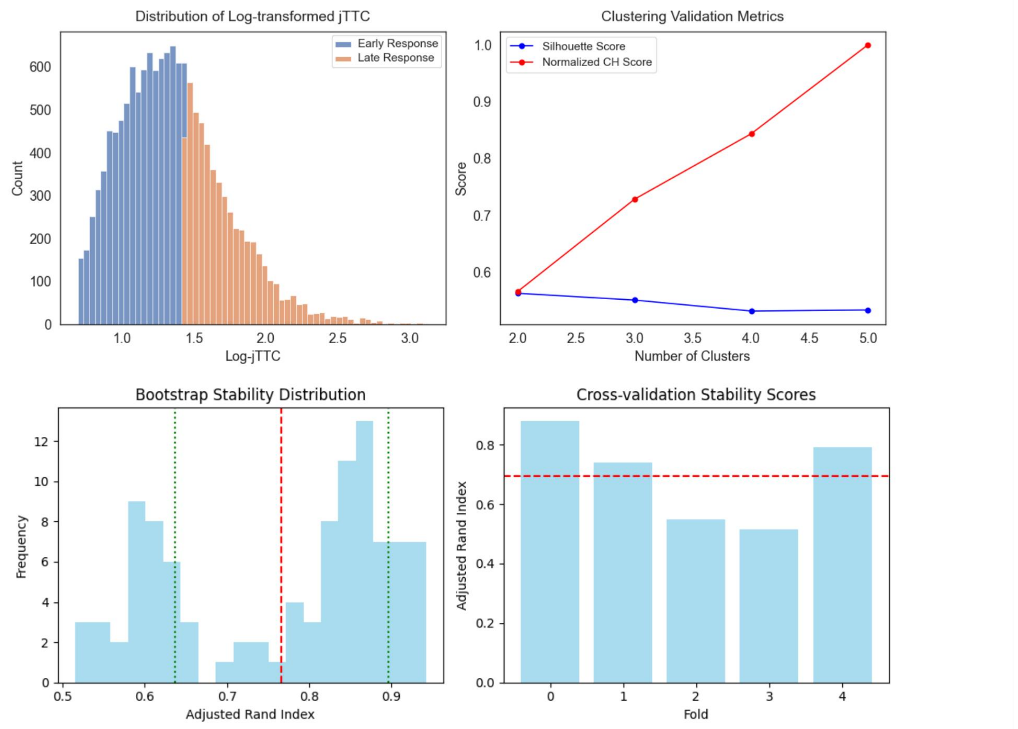


**S4 Fig. Classification and Validation of Temporal-Dynamic Patterns in Pupillary Response.** Top Left: Distribution of log-transformed response time (Log-jTTC), showing two natural groupings of early response (blue) and late response (orange). Top Right: Clustering validation metrics (Silhouette Score and Normalized CH Score) across different numbers of clusters (2-5), showing increasing CH Score with higher cluster numbers. Bottom Left: Bootstrap stability index distribution (red dashed line: mean of 0.753; green dashed lines: ±1 standard deviation). Bottom Right: Five-fold cross-validation stability scores (red dashed line: mean stability of approximately 0.7).

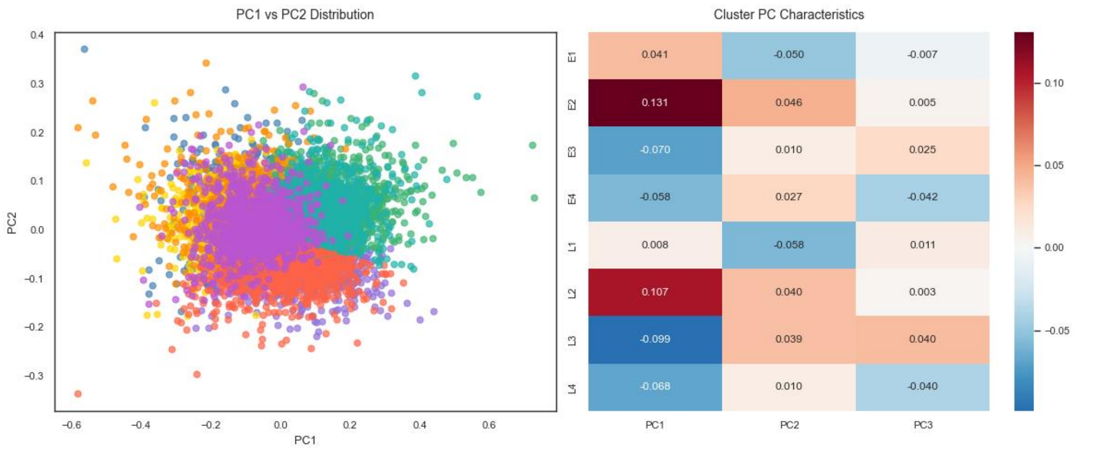


**S5 Fig. Principal Component Analysis of Identified Response Patterns.** Left: Distribution of data points in PC1-PC2 space with color-coded clusters representing different response patterns. Right: Heatmap displaying PC1, PC2, and PC3 values for each of the eight identified patterns (E1-E4: early response patterns; L1-L4: late response patterns), showing principal component characteristics for each cluster.

### Supplementary Results: HMM Analysis

#### Steady State Distribution

Markov chain analysis revealed stable probability distributions of pupillary response patterns across experimental conditions. We conducted chi-square tests and residual analyses on the main effects of Treatment, Sex, Isthreaten, and PSV, with the following results:

Treatment($\chi^2$ (14) = 333.17, $p$ < .001, Cramer's $V$ = 0.107). Residual analysis revealed distinct pattern preferences across treatment conditions. Pattern E1 (PC1+, PC2-, PC3-), characterized by sustained pupil dilation with initial dilation followed by constriction, emerged as a key discriminator between PLC and AVP conditions (PLC: $z$ = 8.55; AVP: $z$ = -9.86). Pattern L3 (PC1-, PC2+, PC3+), showing sustained constriction with enhanced late activation, reached higher equilibrium states in both AVP and LT groups ($z$ = 4.83 and 3.51, respectively) compared to PLC ($z$ = -8.18). Pattern L2 (PC1-, PC2-, PC3+) differentiated AVP from LT, showing elevated equilibrium probability in AVP (z = 3.60) but reduced in LT ($z$ = -2.72).

Sex($\chi^2$ (7) = 250.84, $p$ < .001, Cramer's $V$ = 0.133). Pattern E3 was significantly higher than expected in females but significantly lower than expected in males. Patterns L1 and L2 were significantly higher than expected in males (L1: $z$ = 6.86; L2: $z$ = 2.59) but significantly lower than expected in females (L1: $z$ = -7.27; L2: $z$ = -2.74).

Isthreaten ($\chi^2$ (7) = 73.04, p < .001, Cramer's $V$ = 0.072). Under non-threatening conditions, the system tended toward E1 ($z$ = 3.06) at equilibrium, while threatening stimuli shifted the stable state toward E2 patterns ($z$ = 4.21). Pattern E2 (PC1+, PC2+, PC3+), exhibiting early pupil dilation with enhanced activation, represented a distinct stable state specifically associated with threat processing.

PSV ($\chi^2$ (28) = 549.29, $p$ < .001, Cramer's $V$ = 0.098). Residual analysis revealed systematic velocity-dependent patterns. Slower velocities (V1-V2) predominantly evoked late response patterns (L1, L2), with significant positive residuals for L1 (V1: $z$ = 5.62; V2: $z$ = 4.30) and L2 (V1: $z$ = 5.39; V2: $z$ = 3.89), while inhibiting early patterns, especially E2 (V1: $z$ = -3.68; V2: $z$ = -5.28) and E3 (V1: $z$ = -6.70). Moderate velocity (V3) showed a transitional state with mixed responses, featuring positive residuals for E3 ($z$ = 2.29) and L3 ($z$ = 3.57) but negative for E1 ($z$ = -3.93). Faster velocities (V4-V5) strongly triggered early responses, particularly E3 for V4 ($z$ = 4.56) and both E1 ($z$ = 6.02) and E2 ($z$ = 7.87) for V5, while significantly inhibiting late patterns across L1-L4 clusters. This velocity gradient in pupillary responses likely reflects differential processing of approach speed as a threat cue, with faster approaches demanding more immediate attentional and autonomic resources.

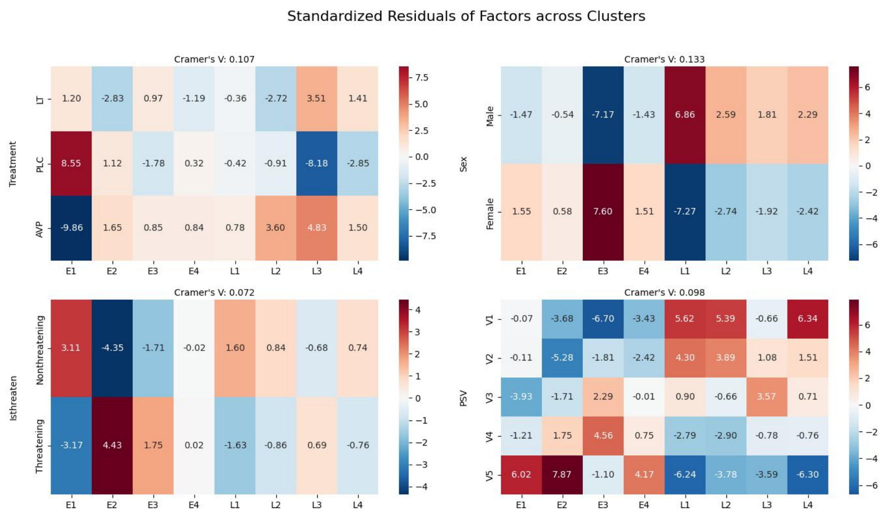


**S6 Fig. Standardized Residuals of Factors across Response Pattern Clusters.** Chi-square and residual analysis examining differential distributions across early (E1-E4) and late (L1-L4) response patterns for: Treatment (top left: LT, PLC, AVP); Sex (top right: Male, Female); IsThreaten (bottom left: Nonthreatening, Threatening); and Physical Stimulus Velocity (PSV) (bottom right: V1-V5). Values represent standardized residuals, with $|z| > 1.96$ indicating significantly higher or lower frequencies than expected by chance. Cramer's $V$ values indicate effect size for each factor.

#### Dynamics Transition

We analyzed transition probabilities using chi-square tests and residual analysis for overall effects, and bootstrap permutation tests (1000 iterations) with FDR correction for specific transitions. This combined approach allowed us to identify statistically significant overall effects while establishing reliable confidence intervals for individual transition probabilities.

##### Treatment

Response patterns showed high self-transition probabilities (0.451-0.630) across all conditions, indicating stable response strategies. Self-transition probabilities varied by treatment: PLC group showed highest stability in E1(0.628) and L1(0.616); LT group exhibited comparable stability in E3(0.609) and E1(0.607); while AVP group was dominated by E3(0.630) and L3(0.583) patterns. All self-transitions were statistically significant ($p$s<.001).

Between-state transitions also showed treatment-specific characteristics. PLC group featured transitions toward E1 (E4→E1: 0.177; E3→E1: 0.175), LT group showed prominent E4→E1(0.166) and E2→E3(0.145) transitions, while AVP group was characterized by E1→E3(0.171) transitions (all $p$s<.001; 95% CI in Table S2).

FDR-corrected group comparisons revealed distinct treatment effects on transition patterns PLC group showed significantly increased probabilities in transitions targeting E1, including E1→E1($z$ = 2.39), E2→E1($z$ = 2.51), E3→E1($z$ = 4.65), and E4→E1($z$ = 2.49). In contrast, AVP group showed reduced transitions toward E1 (E1→E1: $z$ = -4.30; E2→E1: $z$ = -2.83; E4→E1: $z$ = -3.40) but increased transitions involving L2 and L3 patterns (E4→L2: $z$ = 2.92; L3→L3: $z$ = 2.83; L1→L3: $z$ = 2.54). LT group specifically showed reduced transitions toward L2 pattern (E2→L2:

$z = -2.32$; E4→L2: $z = -1.99$).

These results indicate that PLC enhances transitions toward E1 pattern, whereas AVP promotes transitions toward E3, L2, and L3 patterns while reducing E1-directed transitions. Notably, LT selectively inhibits transitions toward L2 pattern (PC1+, PC2+, PC3+), reflecting suppressed activation across all three principal components.

**Isthreaten**

For threat effects, E3→E2 transitions were significantly modulated by Isthreaten ($\chi^2$ (1) = 14.95, $p$-adj = 0.007), with increased probability under threatening conditions ($z$=2.54) and decreased probability under non-threatening conditions ($z$=-2.62).

**Sex**

Sex significantly influenced pupillary response patterns ($\chi^2$ (7) = 250.84, $p < .001$). Male participants showed higher probabilities for transitions involving late response(L-pattern), particularly with elevated L1 ($z = 6.86, p < .001$), L2 ($z = 2.59, p < .01$), and L4 ($z = 2.29, p < .05$) representations. In contrast, female participants exhibited significantly increased E3 pattern ($z = 7.60, p < .001$) with corresponding decreases in late response patterns: L1 ($z = -7.27, p < .001$), L2 ($z = -2.74, p < .01$), and L4 ($z = -2.42, p < .05$).

These sex differences were further reflected in transition probabilities, where males showed higher self-transition stability for L1→L1 (0.611) compared to females (0.547), while females exhibited enhanced E3→E3 stability (0.617 vs. 0.575 in males). Additionally, females demonstrated more frequent transitions from E1→E3 (0.158) and E2→E3 (0.158) compared to males, suggesting sex-specific differences in pupillary response modulation.

**Physical Stimulus Velocity (PSV)**

PSV significantly impacted pupillary dynamics ($\chi^2$ (7) = 73.04, $p < .001$). High-PSV participants showed distinct transition characteristics compared to low-PSV participants. Participants in high-PSV trials exhibited increased self-transition stability in E1→E1 (0.609) and E2→E2 (0.565) patterns, while low-PSV demonstrated enhanced E3→E3 stability (0.631).

Notable differences appeared in steady-state distributions, with high-PSV participants showing greater long-term prevalence of E1 (0.237) and E2 (0.169) patterns, while low-PSV participants showed enhanced E3 representation. This suggests that approaching velocity influences the preferred pupillary response mode, with high-PSV individuals favoring E1 (PC1+, PC2-, PC3-) patterns that reflect specific components of physiological arousal.

**S2 Table. Within-treatment estimated transition probabilities and 95% confidence intervals from the bootstrap permutation test (1000 iterations).**

| Treatment | Transition | Probability | [95% CI] | $p$-adj |
|---|---|---|---|---|
| LT | E3 → E3 | 0.609 | [0.584, 0.632] | ** |
| | E1 → E1 | 0.606 | [0.582, 0.630] | ** |
| | L1 → L1 | 0.563 | [0.531, 0.595] | ** |
| | L3 → L3 | 0.529 | [0.488, 0.568] | ** |
| | L4 → L4 | 0.497 | [0.458, 0.533] | ** |
| | E2 → E2 | 0.493 | [0.455, 0.529] | ** |
| | E4 → E4 | 0.484 | [0.447, 0.516] | ** |

| | | | | |
|---|---|---|---|---|
| | L2 → L2 | 0.473 | [0.430, 0.512] | ** |
| | E4 → E1 | 0.166 | [0.141, 0.190] | ** |
| | E2 → E3 | 0.145 | [0.121, 0.172] | ** |
| PLC | E1 → E1 | 0.628 | [0.607, 0.649] | ** |
| | L1 → L1 | 0.616 | [0.584, 0.646] | ** |
| | E3 → E3 | 0.557 | [0.529, 0.582] | ** |
| | L2 → L2 | 0.511 | [0.471, 0.551] | ** |
| | E2 → E2 | 0.494 | [0.462, 0.527] | ** |
| | E4 → E4 | 0.491 | [0.457, 0.524] | ** |
| | L4 → L4 | 0.467 | [0.428, 0.506] | ** |
| | L3 → L3 | 0.450 | [0.385, 0.512] | ** |
| | E4 → E1 | 0.178 | [0.154, 0.202] | ** |
| | E3 → E1 | 0.175 | [0.156, 0.195] | ** |
| AVP | E3 → E3 | 0.63 | [0.606, 0.653] | ** |
| | L1 → L1 | 0.584 | [0.553, 0.614] | ** |
| | L3 → L3 | 0.583 | [0.545, 0.619] | ** |
| | E1 → E1 | 0.532 | [0.498, 0.564] | ** |
| | E2 → E2 | 0.528 | [0.493, 0.559] | ** |
| | L2 → L2 | 0.522 | [0.488, 0.558] | ** |
| | L4 → L4 | 0.509 | [0.474, 0.545] | ** |
| | E4 → E4 | 0.493 | [0.458, 0.528] | ** |
| | E1 → E3 | 0.172 | [0.150, 0.195] | ** |
| | L4 → L1 | 0.152 | [0.129, 0.177] | ** |

** *p*<.01 after FDR correction.

**S3 Table. Between-treatment differences in transition probabilities and 95% confidence intervals from the bootstrap permutation test (1000 iterations).**

| Treatment | Transition | Diff. | [95% CI] | *p*-adj |
|---|---|---|---|---|
| PLC - LT | E3 → E1 | -0.055 | [-0.080, -0.032] | * |
| | L1 → L3 | 0.045 | [0.026, 0.063] | * |
| PLC - AVP | L3 → L3 | 0.133 | [0.064, 0.203] | ** |
| | E1 → E1 | -0.096 | [-0.135, -0.060] | ** |
| | E3 → E1 | -0.075 | [-0.097, -0.052] | ** |
| | E3 → E3 | 0.073 | [0.039, 0.108] | ** |
| | E4 → E1 | -0.069 | [-0.100, -0.037] | ** |
| | E2 → E1 | -0.060 | [-0.089, -0.031] | ** |
| | L1 → L3 | 0.051 | [0.032, 0.069] | ** |
| | E1 → E3 | 0.039 | [0.013, 0.065] | ** |
| | E4 → E2 | 0.039 | [0.010, 0.068] | * |
| | L3 → E1 | -0.039 | [-0.068, -0.010] | * |
| LT - AVP | E1 → E1 | -0.075 | [-0.115, -0.033] | * |

| | | | | |
|---|---|---|---|---|
| | E4 → E1 | -0.057 | [-0.088, -0.026] | * |
| | E1 → E3 | 0.043 | [0.015, 0.071] | * |
| | L3 → E3 | -0.041 | [-0.066, -0.016] | * |
| | E4 → L2 | 0.023 | [0.007, 0.039] | * |

* *p*<.05 ** *p*<0.01 after FDR correction.

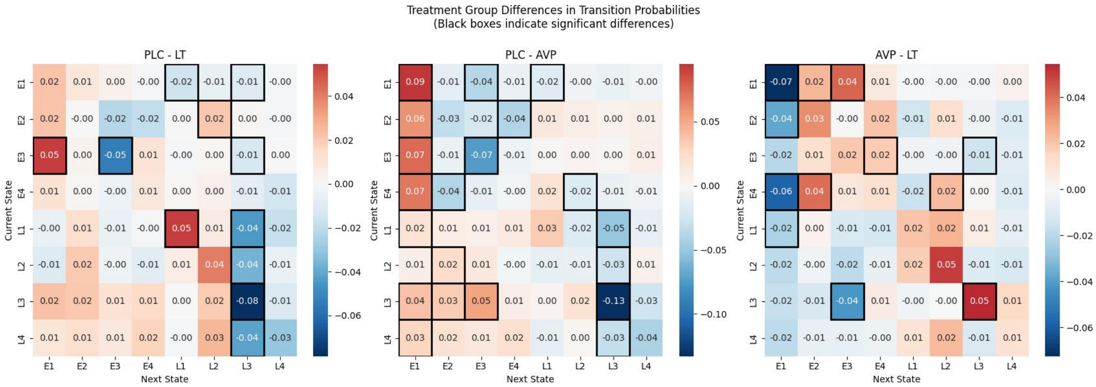


**S7 Fig. Treatment group differences in transition probabilities.** Each matrix shows pairwise comparisons between treatment groups (left: PLC-LT; middle: PLC-AVP; right: AVP-LT). The rows represent current states and columns represent next states. Cell values indicate the difference in transition probabilities between groups (e.g., in PLC-LT matrix, positive/red values indicate higher probabilities in PLC compared to LT group). Black boxes highlight significant transitions (*p*-adj < .05). Color intensity corresponds to the magnitude of probability differences, with red indicating positive differences and blue indicating negative differences. For example, in the PLC-LT comparison, transitions toward L3 state show significantly lower probabilities in PLC compared to LT group, as indicated by blue cells with black borders. Transition probabilities were analyzed using permutation tests (*n*=1000) with FDR correction.

### Supplementary Results: Sex Differences in Patterns

Pattern analysis provides a unifying multi-level account of these differences by identifying distinct sex-biased neurocognitive states:

**Male-biased Profile L1** (+, -, +) is characterized by high sustained vigilance (PC1+) and high cognitive dynamics (PC3+), but low proactive threat preparation (PC2-). This cognitive profile is reflected in distinct pupillary dynamics, showing an initial expansion followed by a pronounced contraction, which aligns with a strategy of generalized environmental monitoring. The L-subgroup (late-response) manifests as longer judged time-to-collision (jTTC), indicating a tendency toward temporal overestimation and delayed responding. Overall, males maintain a high baseline readiness but invest less in specific, sustained threat simulation once a stimulus is categorized, consistent with their lack of post-stimulus threat discrimination. This pattern may reflect a tonic state of exploratory vigilance optimized for broad monitoring rather than deep threat-specific processing (Aston-Jones & Cohen, 2005), which behaviorally manifests as a less precautionary response strategy.

**Female-biased Profile E3** (-, +, +) presents a contrasting pattern: low sustained vigilance (PC1-), high proactive preparation (PC2+) and high internal simulation (PC3+), which marked by pupillary constriction during stimulus approach that transitions to dilation after its offset, suggesting a strategy of targeted resource allocation. The E-subgroup (early-response) was associated with shorter judged TTC, indicating a tendency toward temporal underestimation and accelerated responding. Females appear to suppress initial generalized vigilance to preferentially engage in sustained, internal threat modeling and preparation after stimulus disappearance, directly explaining their robust post-stimulus threat differentiation and behavioral bias towards earlier responses. This suggests a phasic, threat-specific resource investment strategy geared towards detailed evaluation of relevant dangers (Blanchard et al., 2011), which behaviorally manifests as the previously identified precautionary policy of early action.

In summary, the male-biased strategy (Profile L1) manifests as a state of broad, shallow monitoring that maintains generalized environmental readiness at the cost of detailed threat evaluation, resulting in delayed defensive responses. Conversely, the female-biased strategy (Profile E3) facilitates a state of sustained, targeted internal modeling of threats after stimulus offset, which supports superior threat discrimination and promotes the precautionary action of early responses. This functional dissociation provides a parsimonious model that directly links sex-specific neurocognitive states to their distinct behavioral phenotypes.

### Reference


Aston-Jones G., & Cohen J. D. (2005). AN INTEGRATIVE THEORY OF LOCUS COERULEUS-NOREPINEPHRINE FUNCTION: Adaptive gain and optimal performance. *Annual Review of Neuroscience*, *28*(Volume 28, 2005), 403–450. https://doi.org/10.1146/annurev.neuro.28.061604.135709

Blanchard, D. C., Griebel, G., Pobbe, R., & Blanchard, R. J. (2011). Risk assessment as an evolved threat detection and analysis process. *Neuroscience & Biobehavioral Reviews*, *35*(4), 991–998. https://doi.org/10.1016/j.neubiorev.2010.10.016

Dan, E. L., Dînşoreanu, M., & Mureşan, R. C. (2020). Accuracy of six interpolation methods applied on pupil diameter data. *2020 IEEE International Conference on Automation, Quality and Testing, Robotics (AQTR)*, 1–5. https://doi.org/10.1109/AQTR49680.2020.9129915

Eilers, P. H. C., & Marx, B. D. (1996). Flexible smoothing with B-splines and penalties. *Statistical Science*, *11*(2), 89–121. https://doi.org/10.1214/ss/1038425655

Hayes, T. R., & Petrov, A. A. (2016). Mapping and correcting the influence of gaze position on pupil size measurements. *Behavior Research Methods*, *48*(2), 510–527. https://doi.org/10.3758/s13428-015-0588-x

Hershman, R., Henik, A., & Cohen, N. (2018). A novel blink detection method based on pupillometry noise. *Behavior Research Methods*, *50*(1), 107–114. https://doi.org/10.3758/s13428-017-1008-1

Kou, J., Lan, C., Zhang, Y., Wang, Q., Zhou, F., Zhao, Z., Montag, C., Yao, S., Becker, B., & Kendrick, K. M. (2021). In the nose or on the tongue? Contrasting motivational effects of oral and intranasal oxytocin on arousal and reward during social processing. *Translational Psychiatry*, *11*(1), 94. https://doi.org/10.1038/s41398-021-01241-w

Kret, M. E., & De Dreu, C. K. W. (2019). The power of pupil size in establishing trust and reciprocity. *Journal of Experimental Psychology. General*, *148*(8), 1299–1311. https://doi.org/10.1037/xge0000508

Mathôt, S., Fabius, J., Van Heusden, E., & Van der Stigchel, S. (2018). Safe and sensible preprocessing and baseline correction of pupil-size data. *Behavior Research Methods*, *50*(1), 94–106. https://doi.org/10.3758/s13428-017-1007-2

Ramsay, J. O., & Silverman, B. W. (1997). Principal components analysis for functional data. In J. O. Ramsay & B. W. Silverman, *Functional Data Analysis* (pp. 85–109). Springer New York. https://doi.org/10.1007/978-1-4757-7107-7_6

Winn, B., Whitaker, D., Elliott, D. B., & Phillips, N. J. (1994). Factors affecting light-adapted pupil size in normal human subjects. *Investigative Ophthalmology & Visual Science*, *35*(3), 1132–1137.

Zhuang, Q., Zheng, X., Yao, S., Zhao, W., Becker, B., Xu, X., & Kendrick, K. M. (2022). Oral administration of oxytocin, like intranasal administration, decreases top-down social attention. *International Journal of Neuropsychopharmacology*, *25*(11), 912–923. https://doi.org/10.1093/ijnp/pyac059
